# Alterations in the hepatic microenvironment following direct-acting antiviral therapy for chronic hepatitis C

**DOI:** 10.1101/2025.02.17.25321289

**Authors:** Daniel E. Millian, Esteban Arroyave, Timothy G. Wanninger, Santhoshi Krishnan, Daniel Bao, Jared R. Zhang, Arvind Rao, Heidi Spratt, Monique Ferguson, Vincent Chen, Heather L. Stevenson, Omar A. Saldarriaga

## Abstract

**Background and aims.** The first direct-acting antivirals (DAAs) to treat the viral hepatitis C (HCV) became available in 2011. Despite numerous clinical studies of patient outcomes after treatment, few have evaluated changes in the liver microenvironment. Despite achieving sustained virologic response (SVR), patients may still experience adverse outcomes like cirrhosis and hepatocellular carcinoma. By comparing gene and protein expression in liver biopsies collected before and after treatment, we sought to determine whether specific signatures correlated with disease progression and adverse clinical outcomes. **Methods.** Biopsies were collected from 22 patients before and after DAA treatment. We measured ∼770 genes and used multispectral imaging with custom machine learning algorithms to analyze phenotypes of intrahepatic macrophages (CD68, CD14, CD16, MAC387, CD163) and T cells (CD3, CD4, CD8, CD45, FoxP3). **Results.** Before DAA treatment, patients showed two distinct gene expression patterns: one with high pro-inflammatory and antiviral gene expression and another with weaker expression. Patients with adverse outcomes exhibited significantly (p<0.05) more inflammatory activity and had more advanced fibrosis stages in their baseline biopsies than those with liver disease resolution. Patients who achieved SVR had significantly decreased liver enzymes, reduced inflammatory scores, and restored type 1 interferon pathways similar to controls. However, after DAA treatment, patients with persistently high gene expression (67%, pre-hot) still had significantly worse outcomes (p<0.049) despite achieving SVR. A persistent lymphocytic infiltrate was observed in a subset of these patients (76.5%). After therapy, anti-inflammatory macrophages (CD16+, CD16+CD163+, CD16+CD68+) increased, and T cell heterogeneity was more pronounced, showing a predominance of helper and memory T cells (CD3+CD45RO+, CD4+CD45RO+, CD3+CD4+CD45RO+). **Conclusions.** Patients who have more inflamed livers and more advanced fibrosis before DAA treatment should be closely followed for the development of adverse outcomes, even after achieving SVR. We can enhance patient risk stratification by integrating gene and protein expression profiles with clinical data. This could identify those who may benefit from more intensive monitoring or alternative therapeutic approaches, inspiring a new era of personalized patient care.

**Lay Summary:** Direct-acting antiviral (DAA) therapy has dramatically improved the treatment of chronic HCV, making it curable for most people. This study determined gene and protein expression differences in the liver before and after treatment of HCV. These results will lead to a deeper understanding of the changes in the hepatic immune microenvironment with and without the virus present in the liver in hopes of improving patient surveillance, prognosis, and outcome in the future.

## Introduction

Although hepatitis C virus (HCV) infection continues to be a global health challenge, with approximately 50 million infected globally and 1 million new cases each year, the landscape has shifted dramatically in recent years due to the availability of direct-acting antiviral (DAA) therapy^1^. By selectively targeting enzymes essential for viral replication, DAAs have achieved cure rates higher than 90% ^2,3^. This has substantially reduced the number of patients progressing to chronic HCV, the occurrence and recurrence of hepatocellular carcinoma (HCC), and all-cause mortality, which represent a significant milestone in the clinical management of patients infected with HCV ^4,5^. However, despite many patients achieving sustained virologic response (SVR) after DAA treatment, many individuals remain susceptible to reinfection due to the absence of an effective vaccine and the surge in HCV incidence driven by the opioid epidemic. Specifically, injection drug users account for approximately 70% of new HCV infections ^6,7^.

The liver is a unique immune organ enriched with a diverse array of myeloid and lymphoid cells, and it relies on tolerogenic mechanisms to maintain homeostasis ^8^. HCV infection disrupts this balance by triggering the activation of both innate (e.g., type I interferons and IFN-stimulated genes) and adaptive immune responses (e.g., helper and cytotoxic T cells), which play a crucial role in either mediating viral clearance or sustaining chronicity ^9–12^. Inflammatory macrophage activation and cytotoxic T cell responses are typically protective, as they help to control viral replication and eliminate infected cells ^13,14^. However, the impairment of inflammatory macrophages and T cells, the induction of alternatively activated macrophages involved in tissue repair processes, and the promotion of exhausted T cells, characterized by a loss of effector functions and sustained expression of inhibitory receptors, contribute to HCV persistence in the liver ^15–19^. Human immunodeficiency virus (HIV) co-infection exacerbates HCV-induced inflammation and impairs both the innate and adaptive immune responses against HCV ^20^. DAAs have shown the potential to reverse these mechanisms and boost anti-HCV immunity ^21,22^.

In some cases, patients treated with DAAs exhibit persistent inflammation in the portal tracts of the liver, with mild alterations in the fibrosis stage or no improvement ^23,24^. Post-treatment lymphocytic inflammation is distinguished from other types of active hepatitis by its confinement to the portal tracts without interface and lobular activity, and it can still impose diagnostic challenges for clinicians and pathologists managing HCV cases ^25^. Persistent lymphocytic portal inflammation is frequently observed in allograft liver biopsies after treatment, which can be puzzling and lead to uncertainty that SVR has been achieved ^26^. Moreover, it is unclear whether patients who achieve SVR but exhibit histological evidence of persistent lymphocytic inflammation are predisposed to adverse outcomes such as cirrhosis or cancer. Advanced imaging and single-cell platforms have been used to enhance understanding of the changes in the hepatic microenvironment’s cellular composition and spatial distribution ^27,28^. However, studies of the liver after DAA treatment, in the absence of HCV, are limited. This limitation is partly due to the standard of care for HCV, which no longer includes liver biopsy assessment after treatment, thereby reducing the availability of this tissue for analysis ^29^.

This study aimed to analyze the gene and protein expression changes in the hepatic microenvironment in patients before and after DAA treatment for HCV. The study’s uniqueness was the availability of liver biopsies in patients after DAA treatment. This allowed us to correlate individual gene and protein signatures detected in the hepatic microenvironment with clinical outcomes and disease progression.

## Materials and Methods

### Study patients, biopsy collection, and inclusion criteria

We evaluated liver biopsies collected from patients before (i.e., pre-treatment) and after (i.e., post-treatment) DAAs from January 2008 to June 2021. The University of Texas Medical Branch (UTMB) Institutional Review Board approved this clinical study (IRB#13-0511). Written informed consent was obtained from all study patients, and the data collected were de-identified. We previously correlated light microscopic histopathologic findings with clinical outcomes in 10 patients ^25^. Twelve additional patients were added to this cohort (based on tissue availability) to increase the number of patients in each group for molecular and spectral imaging analysis. Sixteen patients had paired pre-and post-treatment biopsies available, while five patients only had pre-treatment biopsies, and one patient had only a post-treatment biopsy collected. We included all ages, women, and minorities in the study. While children were eligible for the study, no participants under 18 met the inclusion criteria at the time of this submission. Demographic data, HCV viral load, and laboratory tests were recorded for each patient included in this study. Liver biopsies were obtained as part of standard care by licensed radiologists and processed in a College of American Pathologists (CAP)-accredited laboratory by licensed histotechnologists as described previously ^25^. The biopsies were evaluated using established numerical scoring systems for histologic features of hepatitis inflammatory activity, fibrosis staging, and steatosis grading ^30–32^. The assessments were conducted by a board-certified liver pathologist (H.S.L) blinded to the patient’s serology, clinical history, and prior pathology reports. The modified hepatitis activity index (MHAI) assesses parameters to evaluate hepatic inflammatory activity, including periportal/periseptal interface hepatitis, confluent necrosis, focal lytic necrosis/apoptosis, and the amount of portal inflammation ^30^. The Ishak criteria were used for staging fibrosis ^30^. Liver biopsy images for each patient were acquired using an Aperio Image Scope Digital slide scanner (Leica Biosystems, Buffalo Grove, IL). Tissue blocks were stored at room temperature and sectioned just before staining whenever possible. Slides that could not be stained within one week were wrapped in parafilm and stored at −80°C with desiccant to minimize moisture exposure.

Liver biopsies used as controls were from patients that met the following criteria: no history of previous HCV infection or any other hepatotropic viral infection, no history of HIV, no history of congenital or acquired liver disease, no history of excessive alcohol or substance abuse, no history of metastatic liver cancer, and no history of immune-mediated liver disease. Their liver biopsies also had to have no histopathologic findings with liver enzymes (alanine aminotransferase (ALT), aspartate aminotransferase (AST), and alkaline phosphatase (ALP) in the normal range at the time of the biopsy.

### DAA treatment regimens and response to therapy

Different DAA combinations were administered for up to 12 weeks to treat the patients according to international guidelines. Patients were retrospectively followed after DAA treatment, and only those who achieved SVR were included. None of the patients had serologic evidence of coinfection with other hepatotropic viruses (e.g., hepatitis A, B, or E) or immune-mediated liver disease at the time of diagnosis or during the study.

### Laboratory tests

We compared laboratory studies ((ALT, AST, ALP), albumin, bilirubin, platelets, prothrombin time (PT), and international normalized ratio (INR)) in the patients pre- and post-DAA treatment (nL=L22). Viral load, genotype, and liver enzymes were measured in blood samples using standard molecular procedures in an in-house CAP and clinical laboratory improvement amendments (CLIA)-accredited clinical chemistry laboratory, as previously described ^25^. Laboratory data were collected for each patient during the baseline liver biopsy, between three- and four months post-DAA treatment, to determine the presence of SVR and at follow-up.

### RNA isolation and quality assurance for downstream applications

RNA was extracted from FFPE unstained sections and fresh liver biopsies and assessed for quality. Samples meeting strict quality criteria (DV300 > 50%, 260/280 ratio ≥ 1.7) were analyzed using NanoString’s PanCancer Immune Panel (NanoString, Bruker Spatial Biology, Seattle, WA, USA).

### nCounter gene expression analysis

The nCounter platform (NanoString, Bruker Spatial Biology, Seattle, WA, USA) is optimized for use FFPE and fresh tissue samples. Studies conducted on specimens of varying ages have shown no significant differences in performance, indicating the platform’s robustness across various sample types and ages ^33^. To determine gene expression profiling, 50 to 150 ng of the isolated RNA were hybridized with the PanCancer Immune Panel, which includes 730 immuno-oncology-related targets and ∼40 housekeeping genes. This panel was chosen since it contains most of the immunological, inflammatory, and anti-viral pathways activated during an HCV infection. The raw data (RCC files) were imported into nSolver™ analysis software v4.0 (NanoString, Bruker Spatial Biology, Seattle, WA, USA). Preliminary analysis evaluated quality control parameters, including imagen QC, binding density, detection limit, and positive controls. Samples with values outside normal ranges (flagged) were assessed to determine if they needed to be excluded. Housekeeping selection, normalization, differential expression, and immune-related scores calculation were performed in the advanced analysis module within the nSolver™ Analysis Software.

### Differential gene expression

Normalized data from the nSolver™ software was used to establish gene expression signatures. To explore the gene expression patterns among HCV pre- and post-DAA therapy groups and liver controls, a hierarchical clustering analysis based on Z-score transformed data was performed using the R statistical environment (version 3.3.2). The results of these analyses were visually represented through heat maps, accompanied by a dendrogram tree. Next, we conducted a principal component analysis (PCA) on the covariance matrix using the regularized log2-transformed gene data in GraphPad Prism v.10 (GraphPad Software, San Diego, CA, USA). A Venn diagram was also constructed using the significant differentially expressed genes (DEGs) between the individual HCV groups and the liver controls. The selection of significant DEGs was based on an adjusted Benjamini-Yekutieli (BY) p-value cut-off <0.01 and a log2 fold change ≤-0.6 or ≥0.6.

The volcano plots in DEGs were used to illustrate gene profiling among HCV pre- and post-DAA therapy groups compared to liver controls and between subclusters. Volcano plots were generated using the nSolver software and visualized using GraphPad Prism v.10 (GraphPad Software, San Diego, CA, USA). Each gene’s −log10(p-value) was plotted on the x-axis, while the log2 fold change was represented on the y-axis. Genes positioned above the predefined threshold adjusted Benjamini-Yekutieli (BY) p-value cut-off <0.05 and log2 fold change ≤-0.6 or ≥0.6) were identified as differentially expressed. The Database for Annotation, Visualization, and Integrated Discovery (DAVID) is a powerful bioinformatics tool and web-based platform that offers a suite of enrichment analysis tools, enabling explore the functional implications of DEGs within the context of biological pathways ^34^. The DAVID tool, integrated with the Reactome database, was used to identify enriched pathways in DEGs among the HCV pre- and post-DAA therapy groups compared to liver controls.

We conducted a cell deconvolution analysis utilizing the online tool CIBERSORT (Cell Type Identification by Estimating Relative Samples of RNA Transcripts) to assess the proportions of immune cell subtypes in different HCV groups and controls. The CIBERSORT tool was employed with default parameters, using the LM22 expression profile matrix encompassing 22 types of immune cells ^35^. We examined each sample’s normalized gene expression levels (log2) of lineage markers and calculated the p-values using 100 permutations.

### Multispectral imaging analysis

Using two additional unstained slides from each tissue FFPE tissue block, we stained with two separate multiplex antibody panels, one for macrophages (CD68, CD163, Mac387, CD14, and CD16) and DAPI and another for T cells (CD3, CD4, CD8, CD45RO, and FoxP3) and DAPI. Staining was conducted manually or with the Ventana Discovery Ultra (Roche Diagnostics, Indianapolis, IN) as previously described ^36^ and they were using the conditions as detailed in **Table S1-2**. Images were acquired using the Vectra 3 Quantitative Pathology Imaging system (Akoya Biosciences, Marlborough, MA). Regions of interest (ROIs) were obtained from at least 50% of the surface area of each liver biopsy from each patient. Stamped areas for acquisition were 2 x 2 (1338 µm x 1000 µm) at a resolution of 0.5 µm (20X). Acquired images from the macrophage panel staining were analyzed using phenotyping algorithms (inForm, Akoya Biosciences) and merged cell seq_files. Multi-component TIFF images from the macrophage and T cell panels were imported into Visiopharm software (Visiopharm, Hoersholm, Denmark). The images were stitched and analyzed using custom artificial intelligence (AI) applications and deep learning algorithms designed for tissue detection, nuclear detection, and cell phenotyping. Phenotypes were calculated as the number of cells per analyzed area across all ROIs collected from each patient. The phenotyping workflow tool was used to guide and annotate each of the patient’s whole-slide images before batch analysis. These customizations added another layer of scrutiny, optimizing the algorithm to identify each phenotype according to the variability of each patient’s collected images.

### Statistical analyses

Data analysis and boxplot figures were performed using GraphPad Prism v10 (GraphPad Software, San Diego, CA, USA). Descriptive statistics were calculated to summarize the mean, median, and standard deviation (SD). Normal distribution was determined using the D’Agostino-Pearson test or the Shapiro-Wilk test. An unpaired t-test (parametric) or Mann-Whitney test (nonparametric), as appropriate, was used to compare the differences between the two groups. A one-way ANOVA with multiple comparisons, Tukey’s post-hoc test (parametric) or Dunn’s (nonparametric), was used to compare the differences between three or more groups. A p-value <0.05 was considered statistical significance. The Fisher test determined the association between gene expression pre-DAA treatment and outcomes. OR ratios >1 are regarded as a positive association. R2 values>0 suggest a positive correlation.

## Results

### Demographics, study design, histological scoring, and clinical data

The main goal of this study was to analyze changes in gene and protein expression in the hepatic microenvironment before and after DAA treatment to determine how the immune landscape responds when the virus has been cleared. We also correlated gene and protein signatures with inflammatory activity, fibrosis stages, and clinical outcomes. We previously correlated the light microscopy findings with clinical outcomes in 10 patients ^25^. In this study, we increased the number of patients to 22 and further evaluated gene and protein expression using nCounter and multispectral imaging. Patient demographics, histological scoring, and clinical data, including viral loads and genotypes, are shown in **Table 1**. The study group consisted of male (14/22) and female (8/22) patients with a mean age of 52 ±L6.04 SD years when the first biopsy was collected and a mean age of 55 ±L7.3 SD years when the post-treatment biopsy was collected. Fifty percent of the patients were non-Hispanic whites (11/22), 27.3% were Hispanic whites (6/22), and 27.3% were African American (6/22) (data not shown). Patients had a mean body mass index (BMI) of 28.22 ±L6.09 SD kg/m2 and 29.7 ±L6.97 SD kg/m2 at the time of the first and second biopsies, respectively (p = 0.52). The HCV viral load ranged from 1.15L×L10^4^ to 1.72L×L10^7^ IU/mL before DAA treatment. All patients achieved SVR (i.e., undetectable viral RNA for 12 weeks or more post-treatment) independent of the DAA regimen. The most prevalent genotype was 1a (12/22), followed by 1b (4/22) and 2b (4/22). The experimental workflow is shown in **Fig. 1A**. Liver biopsies were collected from the study patients pre and post-DAA treatment. RNA from Study Patient Set 1 (**Fig. 1A**) was subject to NanoString nCounter gene expression analysis. Unstained liver biopsy sections from Study Patient Set 2 were stained with the macrophage phenotyping panel (CD68, CD14, CD16, CD163, MAC387, and DAPI). In contrast, unstained slides from Study Patient Set 3 were stained with the T cell phenotype panel (CD3, CD4, CD8, FoxP3, CD45RO, and DAPI) and analyzed by multispectral imaging. After treatment, patients had significantly decreased (p<0.001) inflammatory activity in their liver (mean and SD of 4.81 MHAI ±L 2.10 and 2.4 MHAI ±L1.14 at the time of the first and second biopsies), and significantly decreased (p<0.01) transaminase levels (mean and SD of AST before 67 U/L ± 74, and after treatment 28 U/L ± 11.56; mean and SD of ALT before 74 U/L ± 79.64 and after treatment 28.4 U/L ± 11.6). There were no significant differences in the fibrosis stage or ALP levels after treatment (**Fig. 1B**).

**Figure 1.**
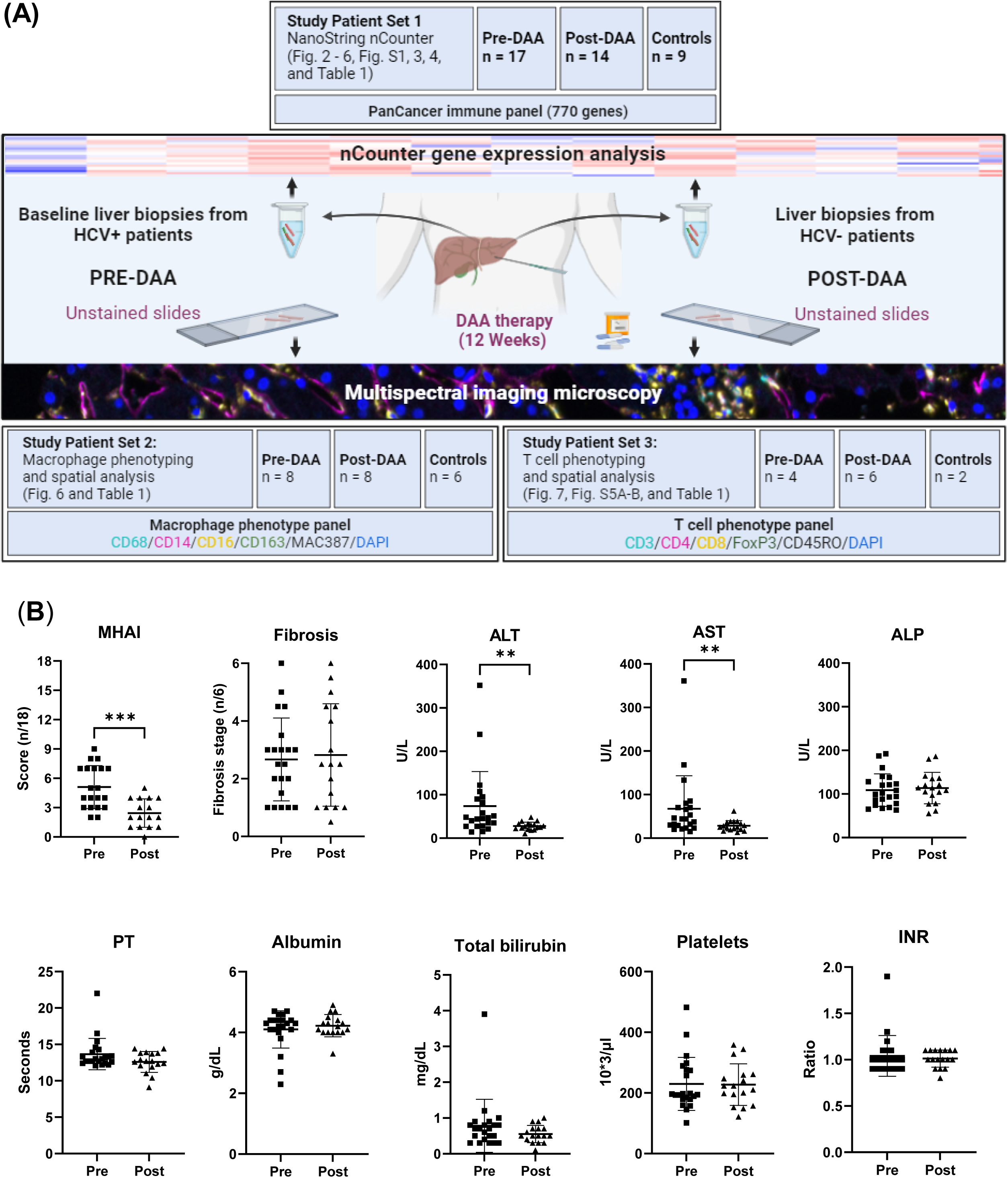
Experimental workflow and comparison of clinical data of patients pre- and post-DAA treatment for viral hepatitis. **C.** (**A**) This study evaluated differences in the hepatic microenvironment of liver biopsies collected from patients with HCV (n = 22), including pre-DAA (n = 21) and post-DAA treatment (n = 17), and controls (n = 15) using gene expression and spectral imaging analysis (see also **Table 1**). The tables with the Study Patient Sets show the number of patients included in each experiment. In the post-treatment group, the biopsy was obtained after achieving SVR. Next, RNA was extracted from fresh liver and FFPE unstained slides for nCounter analysis (**Study Patient Set 1**). Then, spectral imaging was performed by staining pre- and post-treatment liver biopsies of each patient with a macrophage (**Study Patient Set 2**) and T cell (**Study Patient Set 3**) panels and scanned with the Vectra 3 automated quantitative pathology imaging system. Multicomponent TIFFs were imported into the Visiopharm digital software platform to generate phenotype profile maps. (**B**) After DAA treatment, patients had significantly decreased inflammatory activity by liver biopsy evaluation and decreased hepatic transaminase levels (ALT and AST) when compared to pre-treatment. Liver biopsies were graded for inflammatory activity using the MHAI scoring criteria (0-18) and staged for fibrosis using Ishak (1-6) criteria. Normal distribution was tested, and an unpaired t-test was used to compare the differences between the groups. *p < 0.05; **p < 0.01; ***p < 0.001. Abbreviations: Pre, (Pre-DAA treatment); Post, (Post-DAA treatment); MHAI, modified hepatic activity index; ALT, Alanine transaminase; AST, Aspartate aminotransferase; ALP, Alkaline phosphatase; FFPE, Formalin-fixed paraffin-embedded; PT, Prothrombin time; INR, International Normalized Ratio.

### DAA treatment attenuates pro-inflammatory and antiviral pathways in patients with HCV after they achieve SVR

Given that RNA was extracted from FFPE and fresh tissue, we included another step to compare and analyze housekeeping gene expression, excluding samples flagged by the nCounter platform (data not shown). Gene expression patterns formed two main clusters in the heatmap (**Fig. 2A**). One cluster (right) was characterized by overall low gene expression and consisted of controls (A-I, orange labels), most patients post-DAA treatment (brown labels; pt 1, 5, 7-11, 13, 19-21), and some patients pre-treatment (turquoise labels; pt 5-8, 15-18). The second cluster (left) displayed high gene expression levels across the 730 genes analyzed and consisted of most patients pre-DAA treatment (pt 10-14 and 19-22) and three patients post-DAA treatment (pt 6,12, 22). PCA analysis showed a similar clustering pattern (**Fig. 2B**). We then used volcano plots to compare the genes responsible for the heatmap clustering. In agreement with other studies, genes known to be associated with inflammatory (e.g., STAT1, STAT2, CXCL10, CXCL9, IFNAR2) and interferon-induced-antiviral (i.e., ISG15, OAS3, MX1, IFI27, and IFI35) responses were upregulated before DAA treatment when compared to the control (Top) and post-treatment groups (Bottom) (**Fig. 2C, Table S3-4)** ^37,38^. Notably, a significant downregulation of the same genes was observed post-DAA treatment. In most patients who achieved SVR, no significant differences were observed between post-DAA patients and controls (data not shown). Using the most critical genes (as determined by differential gene expression analysis) to compare patients before treatment to controls, we determined that the top enriched pathways were involved in essential biological processes (including genes associated with immunity, innate immunity, and host-virus interactions) and were composed of specific cell components (including membrane, cytoplasm, and secretion systems) (**Fig. 2D**). Similar to other studies, we observed a significant (p<0.01) downregulation of inflammatory (e.g., CXCL10, STAT1) and interferon-induced antiviral genes (e.g., Mx1, OAS3, ISG15/20, IFI27/35, IFIT1) post-DAA treatment ^39^. In addition, we observed heterogeneity in individual patients pre- and post-DAA treatment, as shown by the marked differences in Log2 levels in each group (**Fig. 2E**).

**Figure 2.**
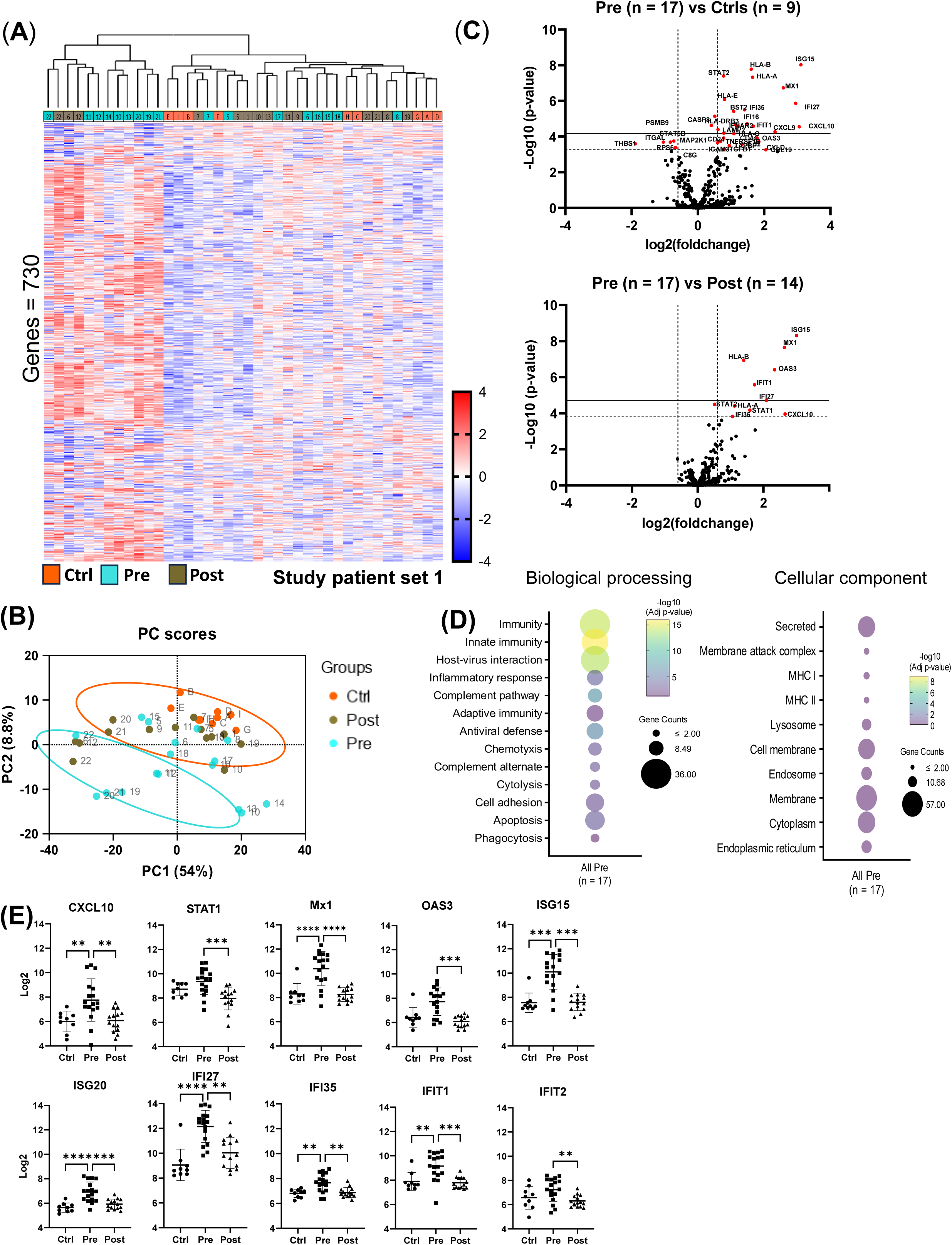
nCounter analysis showed decreased expression of antiviral and pro-inflammatory genes in patients after DAA therapy. (**A**) Cluster analysis using nSolver software categorized gene expression profiling data from liver biopsies of controls (n = 9), patients with HCV pre-(n = 17), and post-DAA treatment (n = 14). The resulting heatmap revealed two main clusters: one was primarily composed of patients before treatment (with increased upregulated genes, red), and the other cluster included mostly patients after treatment, including controls (with more downregulated genes, blue). (**B**) Principal component analysis (PCA) obtained from Log2-transformed data confirmed the clustering observed in the heatmap. (**C**) Volcano plots comparing the gene expression from the liver biopsies of patients pre- and post-DAA treatment vs controls and pre-vs post-DAA treatment show that genes associated with antiviral and pro-inflammatory response were upregulated pre-DAA treatment and returned to baseline post-treatment. Solid and dashed lines indicate the –log10 adjustable (BY) p-value <0.01 or <0.05, respectively. Vertical dashed lines represent the log2 fold change <-0.6 or >0.6. (**D**) Differential gene expression obtained from comparing pre-DAA versus control groups was used to determine enriched pathways using DAVID software and the Reactome database. The y-axis represents the top enriched pathways. The color of the dots represents the −Log10 of the adjusted p-value obtained from the enrichment analysis. –Log10 > 2 (adjusted p-value= ≤0.01) indicates significant enrichment (yellow dots represent higher significance). (**E**) Scatter box plots show significant downregulation of specific antiviral and type I interferon-induced genes in patients post-DAA treatment, like levels observed in the control group. Normal distribution was tested, and One-way ANOVA or Mann-Whitney test was used to compare the differences between the groups. *p < 0.05; **p < 0.01; ***p < 0.001; ****p < 0.0001. Abbreviations: Ctrl (Controls), Pre-(Pre-DAA treatment), Post (Post-DAA treatment).

The relative proportion of immune cells contributing to the liver landscape in the patient groups was estimated using CIBERSORT, which uses a reference dataset of gene expression signatures from known cell types ^35^. While the proportion of monocytes decreased in both the pre- and post-DAA groups, the proportion of pro-inflammatory (M1) macrophages was significantly (p<0.05) increased pre-treatment compared to uninfected controls and decreased (p<0.01) after treatment. T regulatory cells (T regs) were increased before treatment and did not return to baseline after treatment (**Fig. 3A, B**). Specific macrophage and T cell genes accounting for cell deconvolution are shown in **Fig. S1A**. Unexpectedly, gene expression of molecules associated with both inflammatory and anti-inflammatory responses, CD14 and CD16, was significantly elevated in the control group compared to both the pre- and post-treatment groups. At the same time, no significant difference was observed between the pre- and post-treatment groups (**Fig. 3C**). As for molecules associated with specific T cell phenotypes, CD4 and CD8, results were mixed, and the control group exhibited significantly higher expression of CD4 compared to the pre- and post-treatment groups. In contrast, CD8 expression was significantly elevated in both the pre- and post-treatment groups compared to controls (**Fig. 3D**). This dichotomy led us to investigate the protein expression of these markers further, as will be discussed later (see multispectral imaging section).

**Figure 3.**
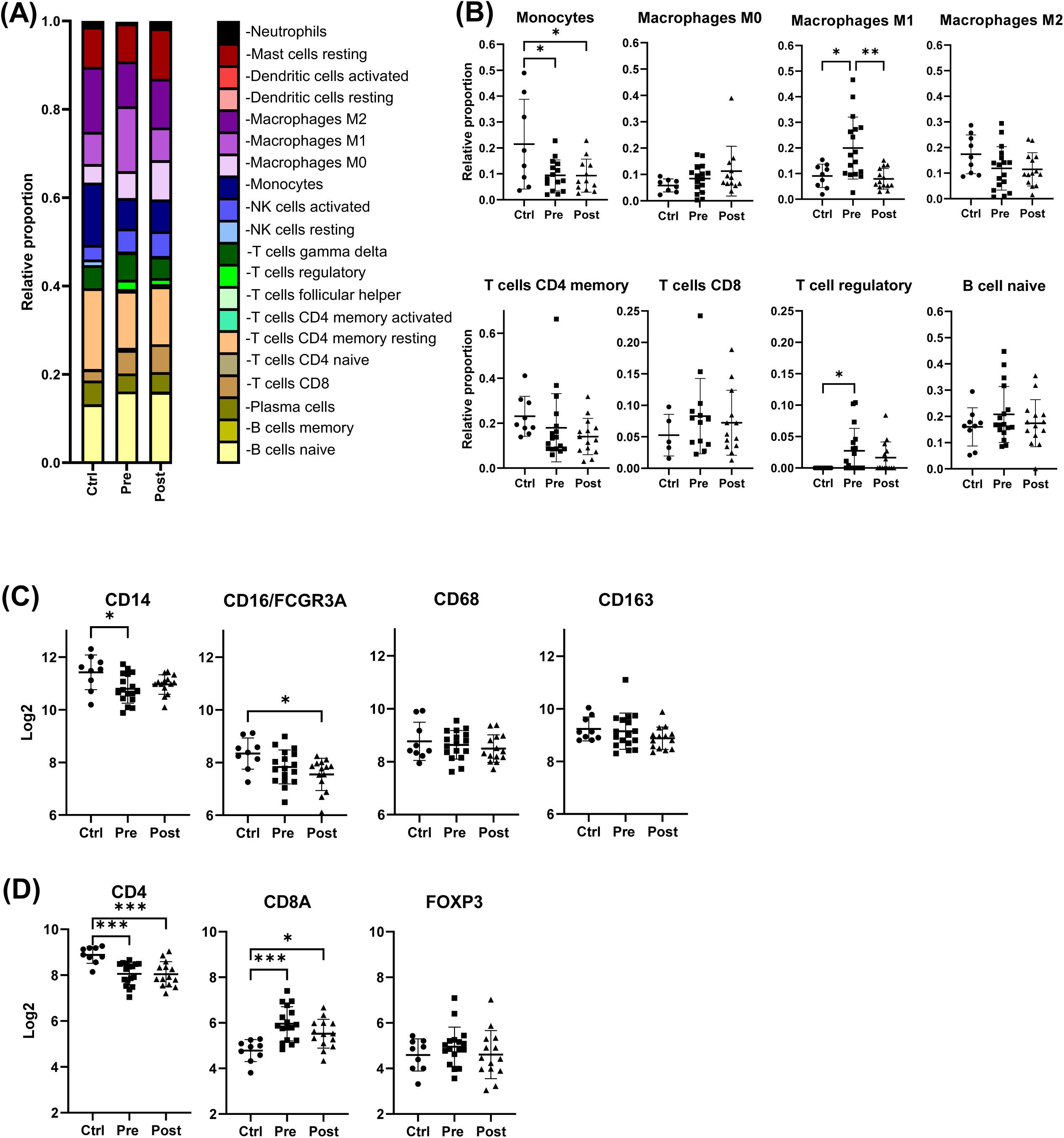
Cell deconvolution analysis showed that pro-inflammatory (M1) macrophages returned to baseline after DAA treatment, while gene expression for regulatory T cells (T regs) remained elevated. (**A**) Gene expression was analyzed using CIBERSORT ^35^. We applied this tool, which uses a reference dataset of gene expression signatures from known cell types, to estimate the relative proportions of different immune cell types in liver biopsies from controls (n = 9) compared with those from patients pre-(n = 17) and post-DAA treatment (n = 14). (**B**) CIBERSORT showed individual variation in specific macrophage and T cell populations among the study groups. After DAA treatment, liver biopsies showed a significantly decreased prevalence of pro-inflammatory (M1) macrophages (light purple), like controls. T reg genes (bright green) significantly increased with HCV infection and were at similar levels after treatment. Specific M1 and T reg genes enriched in the patient’s pre-DAA treatment are shown in **Figure S1A.** (**C-D**) Box plots compare Log2 gene expression of resident, pro-inflammatory and anti-inflammatory macrophages, and T cell markers among the groups. Normal distribution was tested, and a One-way ANOVA or Mann-Whitney test was used to compare the differences between the groups. *p < 0.05; **p < 0.01; ***p < 0.001. Abbreviations: Ctrl (Controls), Pre-(Pre-DAA therapy), Post (Post-DAA therapy).

### A comparison of different clusters identified in the gene expression heat map revealed distinct groups of patients in the pre- and post-DAA groups

We further explored the two clusters identified during the gene expression analysis (**Fig. 2A**). Within the pre- and post-DAA groups, two distinct gene expression and clustering patterns were present (**Fig 4 A-C**). We examined the differences between patients pre-DAA with high gene expression (pre-hot; pt 10-14 and 19-22; n = 9) and low gene expression (pre-cold; pt 5-8 and 15-18; n = 8) and compared each of these groups to controls. Like the pre-DAA group, the pre-hot subcluster had increased inflammatory activity (by MHAI scores) and ALT values (**Fig. S2**), which decreased after treatment (p<0.05). Moreover, pre-hot patients had significantly (p<0.01) increased MHAI scores compared to the pre-cold patients (**Fig. S2**). When compared to controls, patients in the pre-hot group exhibited significantly more upregulated genes with higher fold changes in expression compared to the pre-cold group, as illustrated in the volcano plots and Venn diagram (**Fig. 4A, Fig. S3 - S4, Table S5-S7**). Some genes associated with the JAK/STAT pathway (JAK2, STAT1), TNF (TNFSF4, CYLD), and interleukins (IL-32, IL-17RB) were exclusively expressed in the pre-hot group (**Fig. S4D)**. However, we found that both groups of HCV-infected patients (pre-hot and pre-cold) showed significant gene expression differences (up/downregulated: pre-hot, 162 genes; pre-cold, seven genes; and seven overlapping genes) compared to controls (**Fig. 4C; Fig. S3A, C; Tables S6-7**). Interestingly, differentially expressed genes in the pre-hot patients were not the conventional type I/III IFN-induced genes or antiviral genes, but instead, genes associated with liver cancer progression (i.e., CXCR4, BCL2, FYN) and genes with the potential to exacerbate the inflammatory response (i.e., NLRC5, LTB, CCL3, IKBKE, IRF7, JAK3, TNFAIP3, CXCL9) (**Fig. 4A, Table S5**) ^40–42^. Additional genes found upregulated in the pre-hot subcluster are involved in critical biological pathways, including immunity, innate and adaptive immunity, host-virus interactions, and inflammatory and antiviral responses. They are part of secretion systems (**Fig. 4D-E**). Strikingly, the pre-cold subcluster had conserved pathways associated with antiviral response and innate immunity while showing reduced enrichment of those related to immune and inflammatory responses (**Fig. 4D-E**).

**Figure 4.**
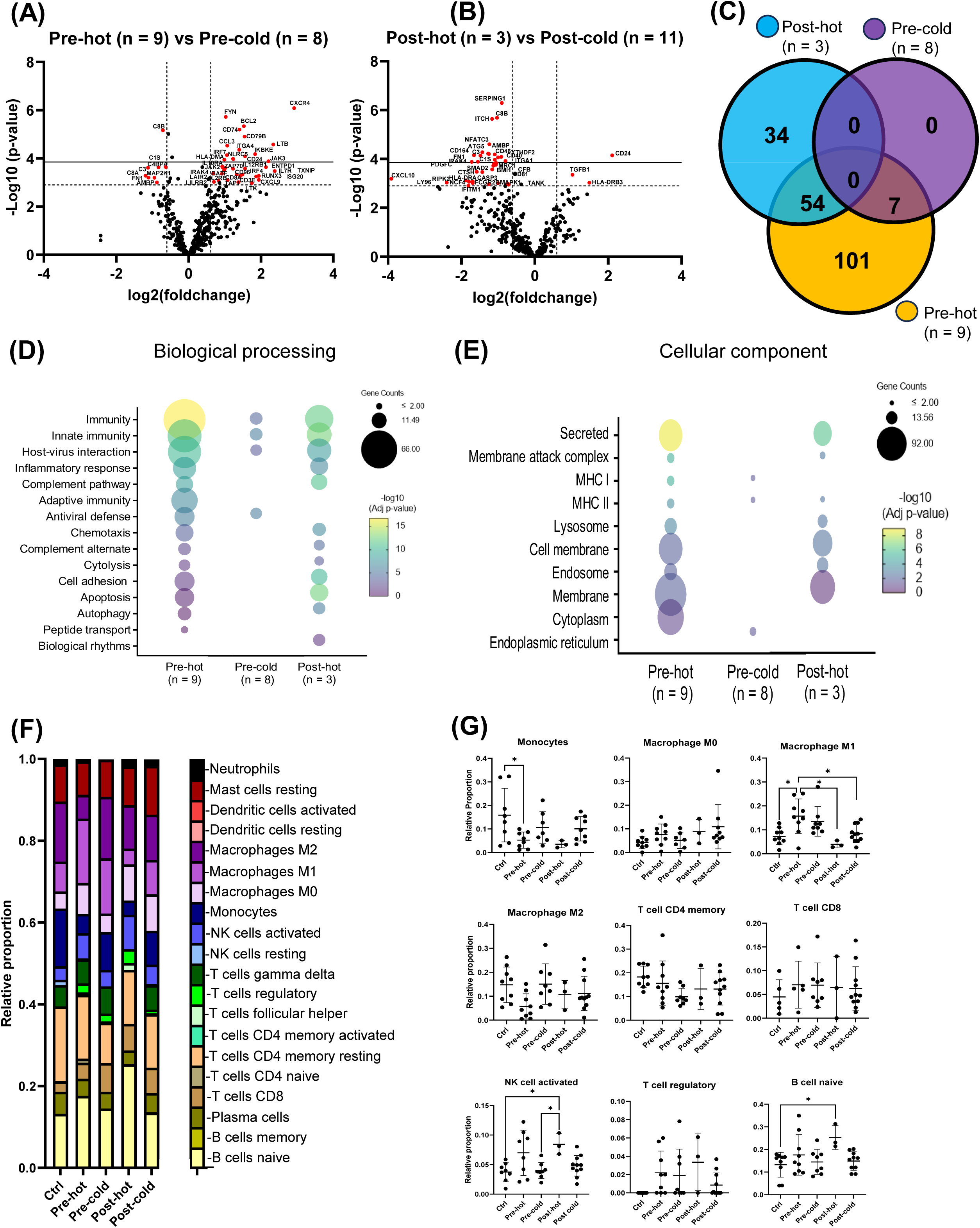
Analysis of gene expression patterns within the pre- and post-DAA groups revealed distinct individual responses to HCV infection and DAA therapy. Within patients infected with HCV and then treated with DAA, we identified different subclusters in the heat map (see **Fig 2A**). Four different groups were compared: the Pre-hot (high gene expression profile Pre-DAA therapy compared to controls), Pre-cold (low gene expression profile Pre-DAA therapy compared to controls), Post-hot (high gene expression Post-DAA therapy comparable to Pre-hot), and Post-cold (low gene expression Post-DAA therapy comparable to controls). Volcano plots were used to compare the gene expression profiles of liver biopsies from (**A**) patients pre-hot versus pre-cold and (**B**) patients post-hot versus post-cold. (**C**) The Venn diagram illustrates significantly expressed genes across pre-cold, pre-hot, and post-hot patients and their overlap. Out of the total 197 differentially expressed genes (the pre-hot expressing 162 genes), 54 overlap between the post-hot and pre-hot, while seven overlap between the pre-cold and pre-hot. In addition, the pre-cold showed a gene expression pattern most like the controls, as it had the fewest significant genes (7 genes) compared to the other groups. (**D-E**) Differential gene expression obtained from comparing pre-hot, pre-cold, and post-hot groups versus liver controls (**see Fig S3-S4 and Table S4-S11**) were used to determine enriched pathways using DAVID software and the Reactome database. The y-axis represents the top enriched pathways. The color of the dots represents the −Log10 of the adjusted p-value obtained from the enrichment analysis. –Log10 > 2 (adjusted p-value= ≤0.01) indicates significant enrichment (yellow dots represent higher significance). Compared to the pre-hot group, the post-hot group was characterized by the activation of persistent immune and inflammatory response pathways without activating antiviral genes and MHC I pathways. (**F-G**) Gene expression was analyzed using CIBERSORT to estimate the relative proportions of different immune cell types in controls and pre- and post-DAA treatment groups (pre-hot, pre-cold, post-hot, and post-cold). Patients pre-hot showed enrichment of M1 cells, and patients post-hot had an increased proportion of naïve B cells (light yellow) and activated NK cells (blue). Specific macrophage (M1 and M2), T and B cells, and NK cells genes enriched in each group are shown in **Fig S1A-B** with corresponding laboratory values in **Fig S2**. Additional Volcano plots showing different comparisons between groups and contributing genes are shown in **Fig S3** and **Table S4-S11**. Normal distribution was tested, and a one-way ANOVA or Mann-Whitney test was used to compare the differences between the groups. *p < 0.05; **p < 0.01; ***p < 0.001.

Similarly, we compared the gene expression patterns of post-DAA patients with high expression (post-hot; pt 6,12,22; n = 3), who clustered with pre-hot patients, to those patients who received DAA and clustered near the control group (post-cold; pt 1, 5, 7-11, 13, 19-21; n = 11) (**Fig. 2B**). Except for TGFβ1 (associated with anti-inflammatory/profibrotic responses) and CD24 (plays a role in cancer proliferation), the other genes in the post-hot patients were significantly downregulated compared with the post-cold group. Cluster analysis of these genes using string indicated the downregulation of complement cascade components (C8B, CFB, C1S, SERPING1, C3, CD46, CD81, CD40) and fibroblast proliferation (PDGFC, BMI1, FN1, SMAD2) (**Fig. 4B, Table S8**). Only the post-hot patients showed significant expression (up/downregulated, 88 genes) compared to controls (**Fig. 4C; Fig. S3B; Fig. S4B; Table S9**). Interestingly, post-hot patients showed an overall downregulation of immune pathways, especially those associated with adaptive immunity, antiviral defense, and MHC I antigen presentation (**Fig. 4D-E**). Significantly regulated genes identified in other comparisons (i.e., pre-hot vs. post-cold patients, post-cold vs. controls) are also shown (**Fig. S3, Tables S10-11).** Specific cell populations were not significantly enriched when comparing the pre-hot to pre-cold or post-hot to post-cold patients; only the pre-hot subcluster shared the same cell pattern initially observed in the pre-DAA treatment group, which consisted of significantly reduced monocytes and increased pro-inflammatory (M1) macrophages that decreased after DAA treatment (**Fig. 4F-G, Fig. 3A, B**). Post-hot patients showed increased NK and naïve B cell activation (**Fig. 4F-G**). The genes responsible for M1 macrophage and B and NK cell enrichment in the pre-hot cluster are shown in the heatmap (**Fig. S1B)**.

Next, we dissected the genes responsible for the identified enriched biological pathways. The heat map in **Fig. 5** highlights and summarizes the activated genes and the associated pathways in each subcluster group compared to the controls. As previously mentioned, the pre-hot cluster generally had more robust gene upregulation than the pre-cold group, which had enrichment of type I/III interferons and anti-viral response pathways. While the pre-hot patients had more than 18 genes (e.g., ISG15/20, CD8A, IRF4, MX1, LTB, CXCL9/10, IFI27, CCL19, CD27/TNFRSF7, SAA1) with a log2 fold change > 2 when compared to controls (**Table S6**), the pre-cold patients only had three genes (ISG15, IFI27, and MX1) (**Table S7**). Interestingly, the log2 ratios of ISG15, IFI27, and MX1 between the two pre-DAA subclusters were 1.32, 1.17, and 1.26, respectively. All the patients post-DAA achieved SVR and had substantially decreased expression of genes related to innate immune and antiviral responses. However, the post-hot patients maintained mixed activation of critical inflammatory (e.g., TNF, TLRs, and NFLB) and anti-inflammatory/profibrotic (e.g., TGFβ, IL4, IL-13, and STAT6) pathways.

**Figure 5.**
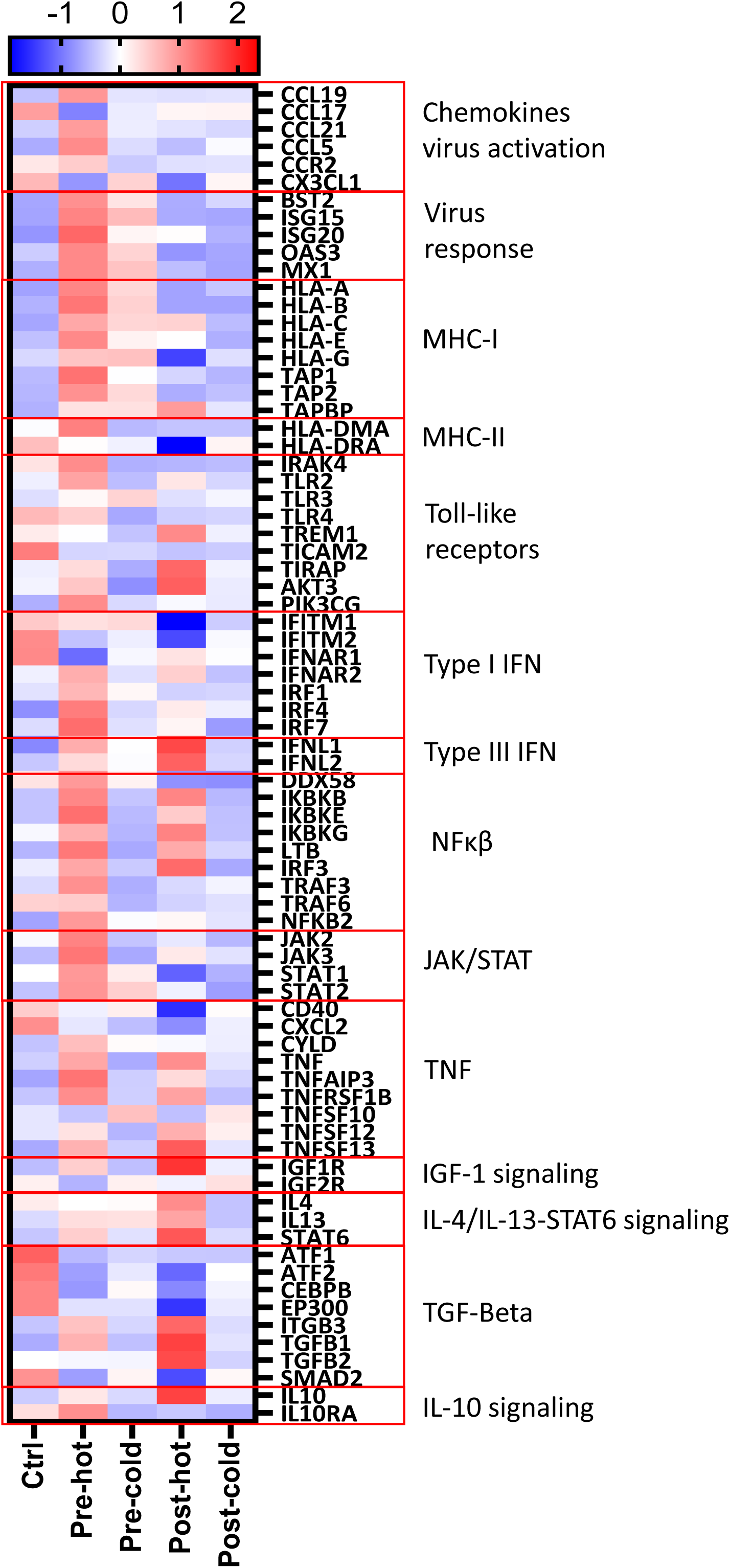
Gene expression patterns reveal distinct inflammatory and fibrogenic signaling pathways despite DAA therapy. We selected differentially expressed genes from **Tables S6-7, 9, 11** (pre-hot, pre-cold, post-hot, and post-cold groups compared to controls). We conducted a STRING network analysis based on these genes to cluster them according to their dominant functional pathway (lowest false discovery rate (FDR)) using an MCL clustering algorithm with an inflation of 5. Subsequently, we generated a heat map using the average Z-score value (intensity range: ±2) for each group. The red boxes represent the clusters of the genes from the more relevant pathways. Viral response and MHC-I markers were upregulated exclusively in the pre-DAA therapy groups (Pre-hot and Pre-cold). Inflammation signals for both pre-hot and pre-cold were associated with type I interferon and TNF/NFkB signaling pathways, with a lower gene expression in the pre-cold group. Conversely, the post-hot group displayed an upregulation pattern associated with IGF-1, IL-4/IL-13-STAT6 alongside TNF/NFkB signaling. Notably, anti-inflammatory and fibrogenic markers such as TGF-beta 1/2 and ITGB3 were significantly expressed in the post-hot group. Abbreviations: Pre-hot (patients with high gene expression pre-DAA treatment), Pre-Cold (patients with low gene expression pre-DAA treatment), Post-Hot (patients with high gene expression post-DAA treatment), Post-Cold (patients with low expression post-DAA treatment), Ctrl (controls).

### Spatial analysis showed that anti-inflammatory macrophages are the predominant phenotype in the hepatic microenvironment after DAA therapy

At the gene level, inflammatory macrophages (M1) were significantly increased during HCV infection and decreased after DAA treatment. There was marked heterogeneity among the different groups, and some patients had persistent lymphocytic inflammation (MHAI) in their liver biopsies (**Table 1**). To evaluate individual patients and preserve the spatial context, we used spectral imaging to determine the primary macrophage phenotypes in the hepatic microenvironment before and after DAAs at the protein level. Unstained liver biopsies from controls and patients pre- and post-DAA treatment (**Table 1, pt 1-8**; **Fig 1A, Study patient set 2**) were stained with a macrophage multiplex panel (CD68, CD14, CD16, CD163, Mac 387, and DAPI). Representative multiplex images from controls (**Fig. 6A, B**), a patient without persistent inflammation post-treatment (pt 6, MHAI = 0/18) **Fig. 6C, D**), and a patient with persistent inflammation post-treatment (pt 2, MHAI = 4/18; **Fig. 6E, F**) are shown. Consistent with our previous studies in steatotic liver disease and HCV, we observed higher expression of CD14+ phenotypes in the controls, which decreased during infection and even further after DAA treatment (p<0.001) (**Fig. 6G**)^36,43^. A significant (p<0.05) increase in the proportion of resident (CD68+ and CD68+/CD163+) and monocyte-derived, pro-inflammatory macrophages (Mac387+, CD68+/CD14+) was observed before DAA treatment in the hepatic microenvironment (**Fig. 6H**). The monocyte-derived and inflammatory (Mac387+, CD68+/Mac387+) populations persisted post-DAA (**Fig. 6H)**. After DAA treatment and SVR, a significant (p<0.05) increase in anti-inflammatory macrophages was observed when compared to controls (CD16+, CD16+/CD163+, CD16+/CD68+) (**Fig. 6I**).

**Figure 6.**
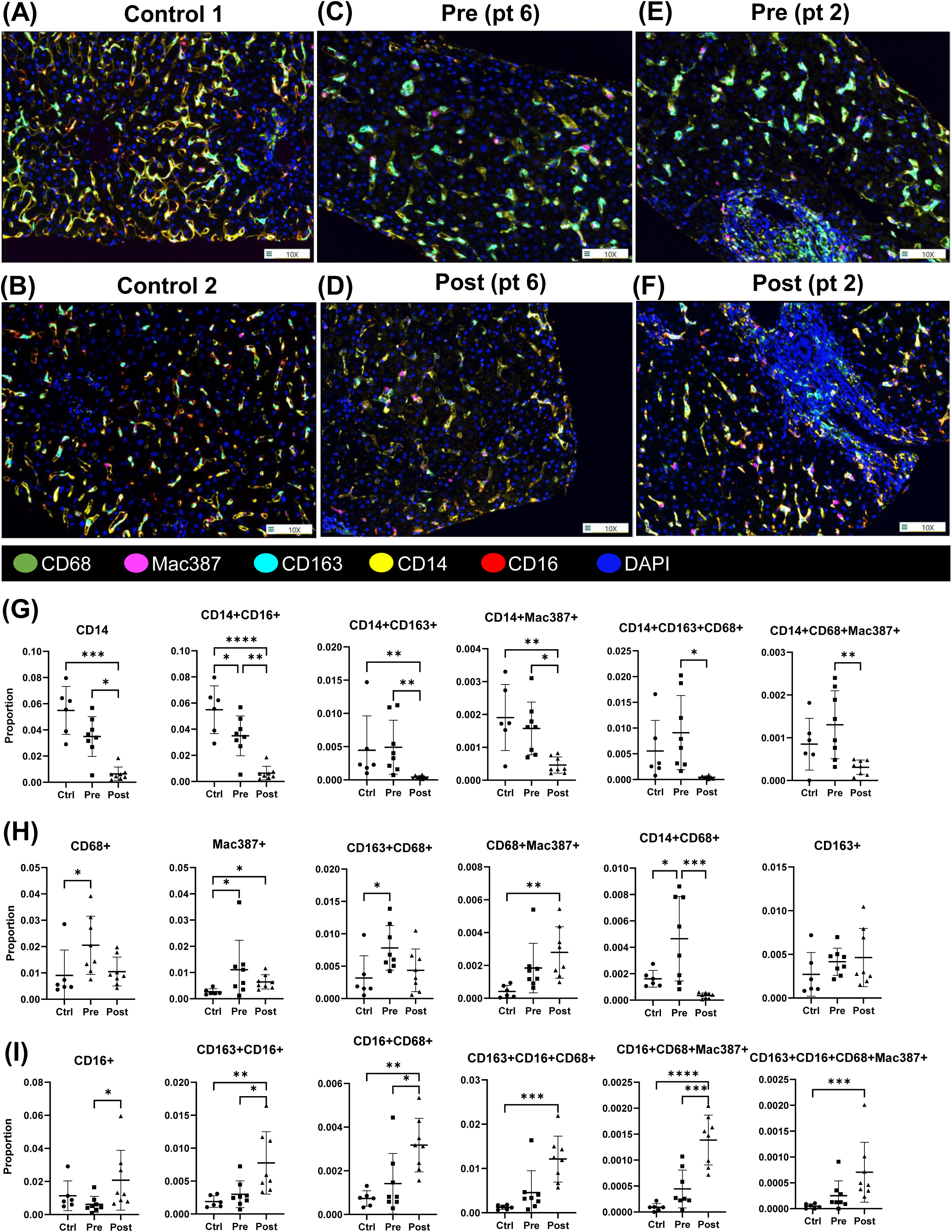
Comparison of macrophage phenotypes in liver biopsies from patients pre- and post-DAA therapy for HCV show a dominant anti-inflammatory response post-DAA. Biopsies were stained with the macrophage multiplex panel (CD68, CD14, CD16, CD163, Mac 387, and DAPI), and images from stained slides were acquired using the Vectra 3 automated quantitative pathology imaging system. Representative multiplex images from controls (**A, B**), a patient (pt 6) without persistent inflammation post-treatment (**C, D),** and a patient (pt 2) with persistent inflammation post-treatment (**E, F**) are shown. (**A, B**) Control patients had more CD14+ cells that decreased with HCV infection and even further post-treatment. (**C, E**) Patients with HCV pre-DAA treatment had increased inflammatory macrophages (CD68+, CD14+, Mac387+, CD68+/Mac387+, and CD68+/CD14+), which decreased post-treatment (**D**), except for pt 2 which had persistent inflammation post-treatment (**F**). Patients with persistent inflammation had increased anti-inflammatory CD16+ and CD16+CD163+ post-treatment. (**G-I**) Shows statistical comparisons of the different phenotypes identified in the control, pre-, and post-treatment groups. Images were acquired at 20x. *Statistical test: non-parametric Wilcoxon rank-sum test, with adj. p<0.05, was used for significance.

### Spatial imaging and t-SNE plots highlighted differences in T cell populations before and after DAA therapy

The presence of marked individual patient heterogeneity in Log2 levels of T cell markers (**Fig. 3**) prompted us to evaluate the patients individually using spectral imaging. We and others have reported the presence of persistent portal lymphocytic inflammation post-DAA treatment despite a significant reduction in the overall inflammatory activity after clearing the HCV with DAAs^23–25,44^. Slides were stained using a multiplex T cell panel (CD3, CD4, CD8, CD45RO, FoxP3, and DAPI; **Fig.1A, Study patient set 3**), and multicomponent TIFF images were uploaded to Visiopharm for analysis using custom-designed algorithms. A detailed phenotype map with the number of positive cells per area shows heterogeneity of T cell phenotypes in individual patients before (pt 5, 8, 12, 13) and after (pt 1, 3, 5, 8, 12, 13) treatment compared to controls is shown (**Fig 7A**). Liver biopsies with an MHAI ≥ 2 (pt 1, 8, 12,13) were considered to have persistent inflammation after treatment, while liver biopsies with an MHAI ≤ 1 (pt 3, 5) were considered not to have persistent inflammation (see **Table 1**). Before and after treatment, there was marked heterogeneity in T cell phenotypes (**Fig. 7A**). CD8+ phenotypes were more frequent in the patients pre-DAA (i.e., CD8+, CD3+/CD8+, CD8+/CD45RO+, CD3+/CD8+/CD45RO+). Infiltration of some of these T cell phenotypes (e.g., CD3+CD8+) agreed with the increased expression of the CD8A gene observed in patients before treatment (see **Fig. 3D)**. Helper T cells (CD3+/CD4+) were also observed pre-DAA. Still, they were more frequent in patients post-DAA treatment (i.e., CD4+, CD3+/CD4+, CD4+/CD45RO+, CD3+/CD4+/CD45RO+) (**Fig. 7A**). Patient 13 had the highest inflammatory activity score before DAAs (MHAI: 5/18; Fibrosis stage: 5/6) and had the highest prevalence of cytotoxic and regulatory T cell phenotypes (e.g., CD3+/CD8+, CD3+/CD8+/CD45RO+, CD3+/CD8+/FoxP3+). The prevalence of these phenotypes remained high after treatment, even though light microscopy appeared to show an absence of persistent inflammation. The patient (pt 1, MHAI: 5/18; Fibrosis stage: 2-3/6) with the highest MHAI after DAAs had increased memory (i.e., CD3+, CD45RO+, CD3/CD45RO+) and helper T cell (i.e., CD3+/CD4+ and CD3+/CD4+/CD45RO+) phenotypes. CD4+ phenotypes were also substantially increased in patients with persistent inflammation (pt 1, 8; **Fig. 7A**).

**Figure 7.**
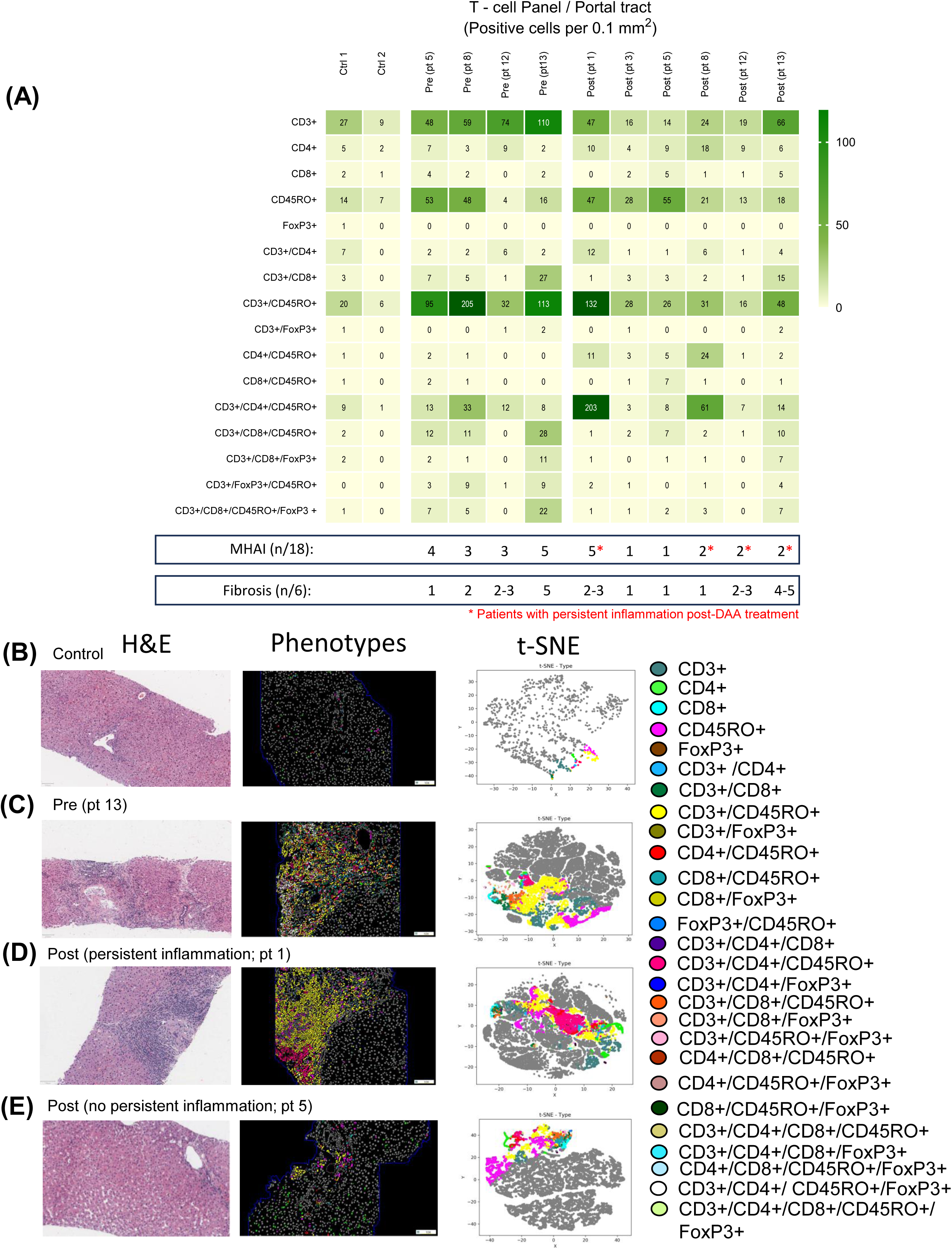
Analysis of T cell phenotypes in liver biopsies from patients pre- and post-DAA therapy for HCV shows heterogeneity between individual patients. (**A**) Unstained slides were stained with the multiplex T cell panel (CD3+, CD4+, CD8+, CD45RO+, FoxP3+, and DAPI). Multicomponent TIFF images were analyzed with Visiopharm using optimized custom AI applications to determine the type and number of T cell phenotypes in the hepatic microenvironment (controls, n = 2; patients with HCV pre-DAA, n = 4; and post-DAA, n = 6). **(A)** The number of phenotypes identified in individual controls and patients with HCV pre- and post-DAA are shown. Variable enrichment of T cells (CD3+), memory T cells (CD45RO+), and cytotoxic T cells (CD3+/CD8+) were observed in patients pre-DAA (pt 5, 8, 12, 13) compared to patients post-DAA treatment with (pt 1,8,12,13; MHAI > 2) and without (pt 3, 5; MHAI < 1) persistent inflammation. (**B-E**) Representative H&E images, phenotypes, and t-SNE plots from controls, patients with HCV pre-DAA, and patients post-DAA with and without persistent inflammation are shown. H&E images were used to evaluate MHAI and fibrosis stages. As illustrated in the phenotype images and tSNE plots, the controls and patients post-DAA therapy without persistent inflammation (**B**, **E**) had low infiltration and diversity of T cell phenotypes compared to patients with HCV pre-DAA therapy (**C**) and post-DAA with persistent inflammation (**D**) (i.e., chronic HCV pre-DAA treatment had increased infiltration of CD3+CD45RO+ (yellow) and patients post-DAA with persistent inflammation had increased CD3+CD4+CD45RO+ (dark pink phenotypes). Each colored dot represents a unique cellular phenotype, and gray dots represent negative cells for all the markers evaluated. MHAI: modified hepatitis activity index; AI, artificial intelligence.

Surprisingly, several T cell phenotypes persisted in the hepatic microenvironment even when the biopsy did not appear to have persistent inflammation by light microscopy (pt 5; **Fig. 7A, B; Fig. S5A**). Representative H&Es, phenotype profile maps, and t-SNE cluster plots (where each color represents a different T cell phenotype, and gray dots indicate cells that were negative for all T cell markers tested in the multiplex assay) are shown for comparison (Control; pre-DAA, pt 13), persistent inflammation (post-DAA, pt 1), without persistent inflammation (post-DAA, pt 5; **Fig. 7B-E**). Individual variation in the T cell phenotypes infiltrating the liver microenvironment of controls and patients pre-DAA and post-DAA treatment are shown in **Fig. S5A-B.**

### Patients who had higher baseline gene expression before DAA treatment had worse clinical outcomes after treatment

Two different gene expression patterns were identified in the biopsies at baseline pre-DAA treatment: the pre-hot subcluster (nine out of 17, 52.9%; pt 10-14 and 19-22) with enriched gene expression and the pre-cold subcluster (eight out of 17, 47.1%; pt 5-8 and 15-18) with low gene expression compared to controls (**Fig. 2, 4; Table S5-7**). Except for the MHAI score (p < 0.01) and a trend toward increased fibrosis in the pre-hot subcluster, no statistically significant differences were observed in laboratory test results, viral load, virus genotype, and clinical data between the pre-hot and pre-cold subclusters (**Fig. S2 and Table 1**).

We then explored whether the patients with different gene expression patterns pre-DAA treatment were associated with poor outcomes post-DAA therapy, defined as progression to cirrhosis, development of HCC, or death related to liver failure. We observed a significant association (Fisher test p<0.049) between poor outcomes and patients exhibiting the pre-hot pattern (**Fig. 8A**). Four out of the nine pre-hot patients (44.4%; pt13, 19-20, and 22) developed cirrhosis, and three out of the nine pre hot (33.3%; pt 10, 14, and 19) died from complications of decompensated cirrhosis. Only one patient with the pre-cold pattern (one out of 8, 12.5%; pt 17) experienced a poor outcome (**Table 1**, **Fig. 8A**). The association of the patients with the pre-hot pattern and the poor outcomes is consistent with the enrichment of genes associated with cancer progression and increased inflammatory responses observed in this subcluster (**Fig. 4A, Table S5**).

**Figure 8.**
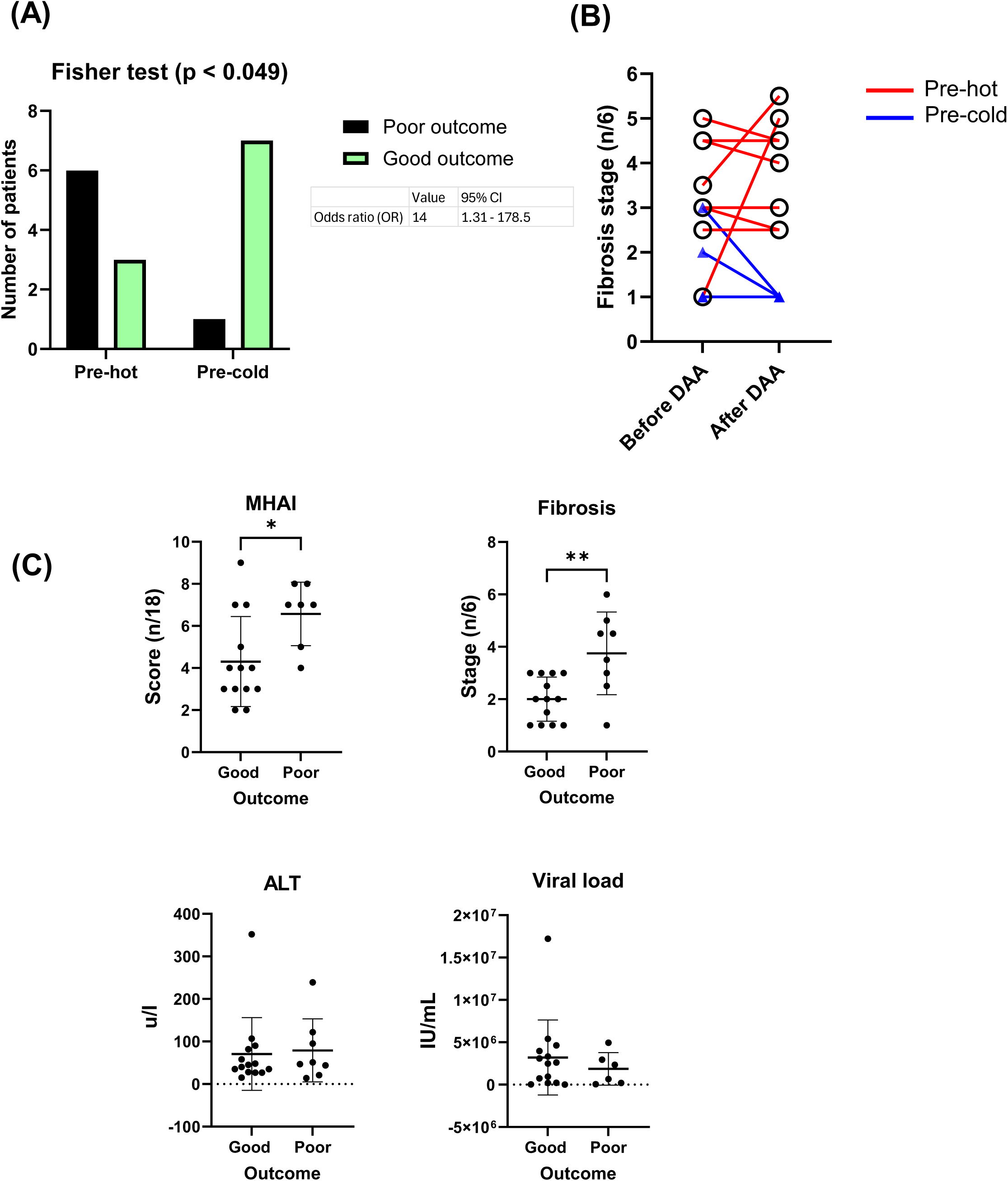
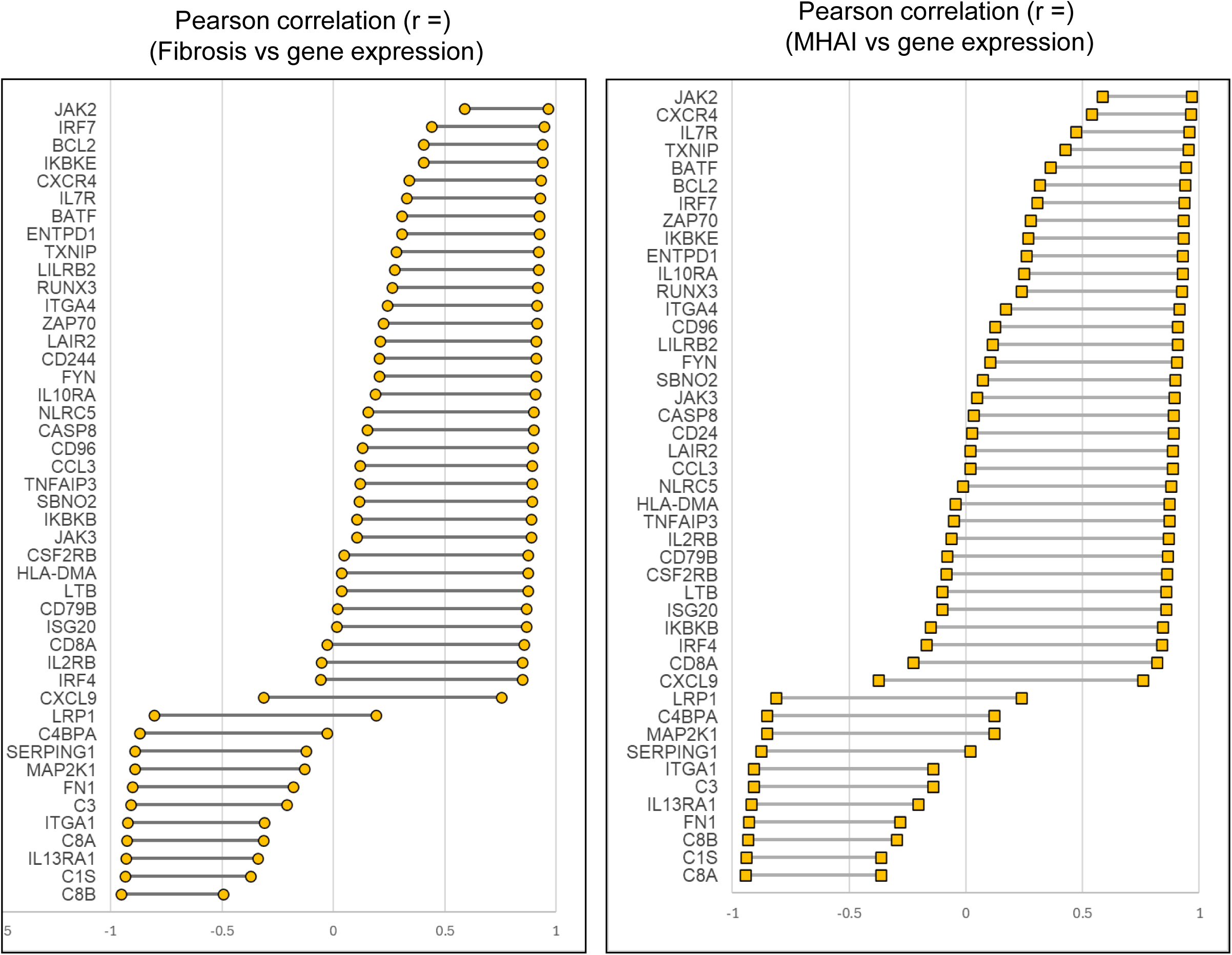
Patients with HCV and increased gene expression show increased poor outcomes and correlated with fibrosis stage post-DAA treatment. **(A)** A positive association of poor outcomes was observed in patients exhibiting the pre-hot pattern (6/9) compared to those with the pre-cold pattern (1/8). (**B**) Patients with increased gene expression (pre-hot; pt 10-13, 19-22) before treatment did not improve their fibrosis post-DAA, while it consistently remained low in the pre-cold group (pt 5-8). (**C**) Patients with poor outcomes had a significantly increased MHAI scores and fibrosis stages compared to patients with good outcomes. (**D**) Genes significantly (p<0.05) increased in the pre-hot subcluster (**Table S5**) compared to the pre-cold group correlated with fibrosis stage and MHI scores after treatment. Abbreviations: MHAI: modified hepatitis activity index. The x-axis represents the r^2^. The Fisher test determined the association between gene expression pre-DAA treatment and outcomes. OR ratios >1 are considered as a positive association. R^2^ values>0 suggest a positive correlation. Normal distribution was tested, and a one-way ANOVA or Mann-Whitney test was used to compare the differences between the groups. *p < 0.05; **p < 0.01; ***p < 0.001.

In most patients treated with DAAs, gene expression returned to control-like levels (11 out of 14 patients; 76%; pt 1, 5, 7-11, 13, 19-21). Only three out of 14 patients (post-hot, 21%; pt 6, 12, 22) showed increased gene expression post-treatment, resembling that of pre-hot patients (**Fig. 4B**, **Fig. 2A**). Two out of these three post-hot patients showed persistent inflammation by histology (pt 12, MHAI: 2; pt22, MHAI: 4) and only one progressed to cirrhosis (Pt 22, **Table 1**).

Among patients with paired liver biopsies (16/22) the fibrosis stage either worsened or remained similar in the pre-hot subcluster (pt 10-13, 19-22), while it consistently remained low in the pre-cold group (pt 5-8) after DAA treatment (**Fig. 8B**). Although patients with poor outcomes (pt 4, 10, 13-14, 17, 19-20, 22) had significantly increased MHAI scores (p<0.05) and fibrosis stages (p<0.01) compared to patients with good outcomes, there were no significant differences in ALT and viral load between the two groups (**Fig. 8C**). We observed that pre-hot patients had 47 genes that were significantly (p<0.05) regulated (up and downregulated) compared to the pre-cold subcluster and mainly consisted of pro-inflammatory genes (**Table S5**). Next, we investigated whether the expression of these genes correlated with inflammatory activity (MHAI) and fibrosis stage after treatment. Thirty of these 47 genes in the pre-hot patients correlated (r = 0–1) with the fibrosis stage, and twenty-two genes correlated (r = 0–1) with the MHAI score (**Fig. 8D**).

Histologic evaluation of liver biopsies showed that 13/17 (76.5%; pt 1-2, 4, 7-8, 10-13, 19-22) had persistent inflammation (MHAI > 2) after DAA treatment, and eight out of these 13 patients (61.5%) started with a pre-hot hepatic gene expression profile before treatment. Only ten (76.9%; pt 7-8, 10-13, 19-22) of these 13 patients had available paired pre- and post-gene expression data (**Table 1**). Six out of these 13 patients (46.2%; pt 4, 10, 13,19-20, 22; MHAI: 2-4/18 and fibrosis stages; 2-6/6) had poor clinical outcomes and five out of these six patients (83.3%: pt 10, 13, 19-20, 22) started with a pre-hot gene expression profile. Two of these 13 patients (15.4%; pt 12, 22) began with a baseline pre-hot gene signature and had a post-hot gene expression signature after treatment. However, these two patients had markedly different clinical outcomes after DAA treatment; one appeared like a control, while the other developed cirrhosis. Of note, patients 4 and 19, who had advanced fibrosis before treatment, developed HCC (**Table 1**). The viral load did not correlate with inflammation scores or fibrotic stages post-DAA treatment. MHAI scores pre-DAA from all patients or MHAI scores from patients with persistent inflammation were not significantly associated with outcomes (Fisher test, p:0.182 and p:0.237, respectively) (data not shown). In addition, six post-cold patients (pt 10-11, 13, 19-21) had a pre-hot signature before DAA therapy. Three of these patients had cirrhosis (pt 13, 19, 20), and two succumbed to cholangiocarcinoma and HCC, respectively (Pt 10,19, **Table 1**). The fibrosis stage alone was unable to predict which patients were more likely to develop a poor prognosis after DAA, as 12 out of 22 (54.5%) patients had a fibrosis stage of 3 or more before treatment, and of those 12, only six (50.0%; Pt 13-14,17,19-20, and 22) developed cirrhosis, cancer and chronic liver-related death (**Table 1**). Patient 20 was the only one who experienced substantial fibrosis progression from stage 1 before treatment to stage 5 after DAA.

## Discussion

HCV is now a curable disease for most individuals^2^. However, despite the successful elimination of the virus, patients are still at risk of liver-related complications, especially if they already have pre-existing liver damage, such as advanced fibrosis ^25^. We and others ^23–25^ have observed that many patients have persistent lymphocytic portal tract inflammation after treatment, and we questioned whether this had any clinical significance. It is well known that macrophages and T cells play essential roles in the protective response to HCV, and chronic infection impairs effective macrophage activation and induces T cell dysfunction ^15,45–47^. Host factors, including male sex, aging, and alcohol consumption, have been shown to have a stronger association with fibrosis progression in chronic HCV than inflammatory activity or virological factors ^48,49^. However, relatively little is known about what happens to the gene signatures or macrophage phenotypes of these immune cells in the liver after DAA. We analyzed liver biopsies from patients before and after treatment to determine if the hepatic microenvironment had residual gene and protein expression changes after the virus was successfully treated. We compared these to liver biopsies from uninfected controls. We analyzed gene expression and characterized intrahepatic immune cells *in situ* using spatial analysis, a technique that preserves the liver architecture ^50^.

All patients with chronic HCV in this cohort achieved SVR and had normal transaminases and reduced inflammatory activity (by MHAI) after treatment (**Table 1**, **Fig. 1)**. However, unlike other studies, we did not observe a significant reduction in fibrosis after achieving SVR (**Fig. 1)**^51,52^. In most patients, the elimination of HCV decreased interferon-mediated inflammatory and antiviral responses to levels like uninfected controls (**Fig. 2, Fig. S3D**). DAA therapy is known to downregulate genes associated with antiviral responses and interferon-stimulated genes (ISGs) in different myeloid and lymphoid cell subsets in the liver ^37,39,53–55^. Kupffer cells have also been shown to be the primary cellular source of hepatic IL-1β during HCV infection ^56^. Studies have shown that this normalization cannot be achieved in patients treated with IFN-based therapies^57^. Correlating with the observed decrease in gene expression of several inflammatory mediators after DAA, macrophages in the liver showed a significant shift from pro-inflammatory (M1-like) to anti-inflammatory (M2-like) phenotypes at the gene and protein level (**Fig. 3, 6**). HCV induced an inflammatory response marked by the upregulation of pro-inflammatory M1 macrophage activation (CD80, CCL5, CXCL9), which decreased post-treatment (**Fig. 3B, Fig. S1A**). Overall gene expression of CD14 and CD16 decreased during infection and post-treatment. In contrast, the expression of CD68 and CD163 remained stable (**Fig. 3C**). A similar analysis of chronic HCV-infected human livers post-DAA treatment showed similar results for CD68 ^52^. Recently, it was demonstrated that the presence of different myeloid subpopulations in the liver, including several CD14+ and CD16+ subsets, were differentially up- or down-regulated post-DAA treatment and contributed to the whole pool of ISG and MHC-II genes expressed during HCV infection ^55^.

The lack of significant changes in the gene expression of CD14, CD16, CD68, and CD163 pre-vs. post-treatment (**Fig. 3C**) may be explained by the differential up/down-regulation of specific subsets post-treatment ^55^. Alternatively, nCounter gene expression analysis was conducted on homogenized liver tissue, which may have diluted the detection of specific cell types. Due to the latter possibility, we moved to spatial imaging to further characterize macrophage changes before and after DAA. With this approach that preserves the liver architecture, we detected an overall shift in pro-inflammatory (i.e., CD14+ macrophage populations) to anti-inflammatory (i.e., CD16+ macrophage populations) cell phenotypes (**Fig. 6G-I**). Chronic HCV infection causes constant inflammatory cytokine production by macrophages, so it makes sense that the elimination of the virus would significantly decrease the proportion of these phenotypes in the liver ^56^. CD16+ phenotypes predominated after treatment and were often at considerably higher proportions than controls (**Fig. 6I**). These observations align with another study that showed that the central CD16+ subpopulation increased following DAA therapy ^55^. Unlike the gene expression results, spatial imaging detected a significant decrease in the M2-like CD14+/CD163+ macrophage phenotype and an increase in the anti-inflammatory/wound-healing CD16+/CD163+ macrophage phenotype after treatment (**Fig. 6 G-I**).

A closer look at the heat map shown in **Figure 2** identified an apparent separation of patients in both the pre- and post-treatment groups with high or low gene expression levels compared to the other patients. Further analysis revealed two distinct inflammatory patterns before treatment. The first pattern (pre-hot) had high expression of genes related to pro-inflammatory (NLRP3, TLR2-4, NFkB, Jak2-3/STAT1-2, TNF) and antiviral (ISG15/20, MX1, OAS3) responses, type I/III IFNs ( IFITM1-2, IFNAR1-2, IRF1,4, IFNL1-2), and MHC-I/II antigen presentation (HLA-C, TAP1-2, HLA-DMA, HLA-DRA). The second pattern (pre-cold) included a similar interferon and antiviral gene profile with a smaller number of genes (ISG15/20, IFI27, MX1, HLA-A/B, STAT2) and lower overall gene expression (**Fig. 2, 4; Tables S5-7**). Although we did not evaluate host genetic variations in this cohort of patients, it is well known that variants of type III IFNs (IFN-λ3/4 gene) influence differential levels of ISG expression, HCV pathogenesis, and response to IFNα and DAAs treatment ^58,59^. Interestingly, by gene expression after achieving SVR, only three patients (post-hot, pt 6, 12, and 22) showed sustained expression of type I IFNs (IFNAR1/2, IRF4), strong expression of type III IFNs (IFNL1-2), and the upregulation of genes and pathways associated with a mixed pro-inflammatory and anti-inflammatory/profibrotic activity (i.e., TLRs, NFkB, TNF, TGFβ, IL4/IL13, IL10) (**Fig. 4B-C**; **Fig. 5**).

Restoration of specific T cells, including cytotoxic and exhausted CD8 T cells, has been observed by isolating these cells from patients’ blood after DAA treatment ^22,60^. In contrast, other systemic T cell populations, like mucosal-associated invariant T (MAIT), are known to remain dysfunctional after DAAs ^61^. Due to these observations and the uncharacterized, persistent portal lymphocytic inflammation after DAA treatment in some patients ^23–25,44^, we analyzed differences in T cell-related genes before and after treatment. In the liver, we observed reduced gene expression of CD4 with chronic HCV infection that did not significantly increase after DAA treatment. This was expected since chronic HCV infection can lead to clonal exhaustion and deletion of HCV-specific CD4 T cells (**Fig. 3D**). Another possible cause for this observation is that several patients were coinfected with HIV ^62–64^. However, there was no significant difference in CD4 gene expression between patients who were and were not coinfected with HIV before and after treatment (**Fig. S6 and Table S12**). Additionally, all patients were on HAART therapy and had similar absolute T cell counts before and after treatment. A comparison of the co-infected patients pre- and post-treatment revealed no significant differences in gene expression (**Fig. S7**). As expected, CD8A expression in the liver increased with chronic HCV infection and trended down after treatment (**Fig. 3D**). However, marked individual patient heterogeneity in Log2 levels prevented statistically significant differences from being detected when comparing the pre- and post-treatment groups. ^65,66^.

Due to the observed heterogeneity in T cell phenotypes in the liver at the gene level, we used multispectral imaging to more clearly compare differences in patients before and after DAA (**Fig. 7A**). Similar to the gene expression results, CD8+ T cells (CD3+CD8+) increased with HCV and persisted in some patients after DAA (**Fig. 3D**, **Fig. 7A**). CD4+ T cell phenotypes became slightly more prevalent after DAA treatment (**Fig. 3D**, **Fig. 7A**). Persistent portal tract inflammation was observed in 76.5% of the study patients after DAA and all of patients had normal transaminases after SVR (**Table S1**). Some studies have reported that about 10% of the patients with ongoing inflammation will still have mildly abnormal liver enzymes after treatment ^44^. As observed in **Figures 3 and 7**, in both the gene and protein levels, patients had marked heterogeneity in the proportions of various T cell phenotypes. This again caused a lack of significance when comparing these cells between the different patient groups (data not shown). Memory T cells (i.e., phenotypes containing CD45RO) were responsible for much of this variability. They persisted after antiviral therapy (**Fig. 7**). These phenotypes were more prevalent in patients with persistent inflammation following SVR (i.e., Pt 1, 8, 12, and 13).

Memory T cells have been shown to inhibit NLRP3 inflammasome activation ^67^, which was upregulated in the pre-hot subpopulation pre-DAA (**Table S6**). Interestingly, we previously demonstrated that CD3+CD45RO+ memory T cells had a stronger correlation with fibrosis progression in patients with metabolic dysfunction associated steatotic liver disease (MASLD/MASH) ^68^. We also compared individual variations in T cell phenotypes infiltrating the liver microenvironment of controls and patients pre- and post-DAA treatment with and without persistent inflammation (**Fig. S5A-B)**.

Next, we compared patient laboratory data, histopathologic features, gene expression, and spectral imaging results to determine if any were correlated with disease progression. Patients with poor clinical outcomes had significantly higher inflammatory activity scores and fibrosis stages before DAA (**Fig. 8C**). Six patients had advanced fibrosis/cirrhosis (27.2%; pt 13-14,17, 19-20, and 22). Two patients with cirrhosis died from ESLD (pts 14 and 20). Out of the 22 patients, three developed cancer and succumbed to the disease (13.6%; pt 4: HCC, pt 10: CCa, and t 19: HCC). However, studies have shown that patients with HCV-related HCC who obtain SVR achieve a 60% - 70% lower risk of 5-year all-cause and liver-related mortality^69^. Interestingly, differentially expressed genes in the pre-hot versus the pre-cold patients showed increased expression of genes associated with liver cancer progression (CXCR4, BCL2, FYN), like other studies ^40–42^. A comparison of the overexpressed genes (including JAK2, IRF7, BCL2, IKBKE, CXCR4) in the pre-hot subcluster (**Fig. 4A**) when compared to the pre-cold subcluster (**Table S5**) had a positive correlation with inflammatory activity scores and fibrosis stages post-DAA treatment (**Fig. 8D and Table 1**). Patients with high gene expression (66.7%, pre-hot) more often had poor clinical outcomes and more advanced liver disease with higher fibrosis stages and inflammatory activity after treatment (**Fig. 4-5, Fig. 8**). Viral load did not correlate with inflammatory activity scores or fibrotic stages after DAA treatment (data not shown) ^24,44^.

A significant strength of this study was the use of liver biopsies obtained both before and after DAA treatment. This allowed us to evaluate histopathologic features, gene and protein expression changes, and individual responses to HCV infection in 22 patients who achieved SVR. Post-treatment liver biopsies are exceedingly rare, allowing us to evaluate the shift in the hepatic microenvironment with and without the virus. We correlated gene and protein signatures with clinical and disease progression, providing valuable insights into the consequences of viral clearance and changes in the immune response after DAA therapy. Unfortunately, some of the paired liver biopsies did not have enough tissue for histology, RNA extraction, and the two spectral imaging panels to be conducted on the same patients. This decreased the sample size of several experiments.

The gene expression analysis identified distinct pre-treatment inflammatory profiles characterized by high (pre-hot) or low (pre-cold) expression of ISGs and antiviral responses. Notably, the pre-hot pattern correlated with adverse outcomes (cirrhosis progression, HCC, and liver failure-related mortality), highlighting the potential for these patterns to serve as prognostic markers. Despite initial variability in gene expression, MHAI scores, and fibrosis stages, DAA treatment effectively controlled viral infection, deactivated antiviral pathways, and returned inflammation to baseline levels in most patients. This study is one of the first to report heterogeneous macrophage subsets in liver biopsies using multiplex immunofluorescence, confirming recent single-cell results that show dynamic and specific gene expression changes in some of these populations post-DAA treatment, underscoring the importance of studies of liver tissue when compared to blood samples. Most persistent inflammation observed in liver biopsies post-treatment appears to be memory T cells. Integrating gene expression profiles with clinical data could enhance risk stratification for patients at risk of poor outcomes, thereby identifying individuals who could benefit from more intensive monitoring and lifestyle modifications to prevent further liver disease progression.

## Data Availability

All data pertinent to this study are either included within the main article or provided as supplementary materials.

## List of abbreviations

HCV: hepatitis C virus
DAA: direct-acting antiviral
SVR: sustained virologic response
POST: post-daa treatment
PRE: pre-daa treatment
HCC: hepatocellular carcinoma
IFN: interferon
HIV: human immunodeficiency virus
UTMB: university of Texas medical branch
IRB: institutional review board
CAP: college of American pathologist
MHAI: modified hepatitis activity index
ALT: alanine aminotransferase
AST: aspartate aminotransferase
ALP: alkaline phosphatase
CLIA: clinical laboratory improvement amendments
FFPE: formalin-fixed, paraffin-embedded
RNA: ribonucleic acid
RCC: resource compiler files
PCA: principal component analysis
DEG: differentially expressed genes
ROIs: regions of interest
ANOVA: analysis of variance
DAPI: 4′,6-diamidino-2-phenylindole
JAK: janus kinase
STAT: signal transducer and activation of transcription
BMI: body mass index
TNF: tumor necrosis factor
TGFB: transforming growth factor beta
MHC: major histocompatibility complex
NKC: natural killer cells
ISG: interferon-stimulated genes
IFI: interferon inducible protein
MX: mixovirus resistance
TIFF: tagged image file format
H&E: hematoxylin and eosin
TSNE: t-distributed stochastic neighbor embedding
MAIT: mucosal-associated invariant T cells
HAART: highly active antiretroviral therapy
NLRP: nucleotide-binding domain, leucine-rich-containing family, pyrin domain containing
MASLD: metabolic dysfunction-associated steatotic liver disease
MASH: metabolic dysfunction-associated steatohepatitis
ESLD: end-stage liver disease
PT: prothrombin time
INR: international normalized ratio
BY: Benjamini-Yekutieli
STRING: search tool for the retrieval of interacting genes/proteins
FDR: false discovery rates
MCL: Markov cluster algorithm
pt: patient
AI: artificial intelligence

## Acknowledgments

We would like to thank the UTMB Institutional Biorepository who provided research histology services in support of this study.

## Author contributions

Data curation: DEM, EA, OAS, JZ. Formal analysis: DEM, EA, OAS, TGW, SK, AR, HS. Methodology: DEM, EA, OAS. Software: TGW, DB, SK, AR. Validation: EA, OAS. Resources: MF. Project administration, Conceptualization, Supervision: HLS. Writing – original draft: OAS, DEM. Writing – review & editing: DEM, HLS, OAS. Reviewed the manuscript: all authors.

## Data availability statement

All data relevant to the study are included in the article or supplementary material.

## Supplementary Figure Legends

**Supplementary Figure 1.** Genes included in the PanCancer panel, utilized by the CIBERSORT tool to estimate the abundance of macrophages, T cells, and NK cell populations, were analyzed: (**A**) before and after DAA treatment and (**B**) within pre- and post-treatment subclusters.

**Supplementary Figure 2. Comparison of clinical data from patients pre- and post-DAA treatment subclustered by gene expression patterns.** We compared the clinical and laboratory data in patients pre- and post-DAA therapy with two distinct patterns of gene expression and clustering: Pre-hot (high gene expression profile pre-DAA therapy compared to controls), pre-cold (low gene expression profile pre-DAA therapy compared to controls), post-hot (high gene expression post-DAA therapy comparable to pre-hot), and post-cold (low gene expression post-DAA therapy comparable to controls). The pre-hot subcluster showed an increased MHAI score and ALT, which decreased post-DAA treatment. Liver biopsies were graded for inflammatory activity using the MHAI scoring criteria (0-18) and staged for fibrosis using Ishak (1-6) criteria^30^. Normal distribution was tested, and an unpaired t-test was used to compare the differences between the groups. *p < 0.05; **p < 0.01; ***p < 0.001. Abbreviations: Pre, (pre-DAA treatment); Post, (post-DAA treatment); MHAI, modified hepatic activity index; ALT, Alanine transaminase; AST, Aspartate aminotransferase; ALP, Alkaline phosphatase; FFPE, Formalin-fixed paraffin-embedded; PT, Prothrombin time; INR, International Normalized Ratio.

**Supplementary Figure 3.** Differentially expressed genes from patients pre- and post-DAA treatment. The volcano plots were constructed using fold-change values and adjustable (BY) p-values. The vertical lines correspond to 0.6-fold changes up and down, and the solid and dashed horizontal lines represent an adjustable (BY) p-value <0.01 or <0.05, respectively. The red dots in the plot represent the differentially expressed mRNAs with statistical significance. Abbreviations: Ctrls, controls; Pre-hot (high gene expression profile pre-DAA therapy), pre-cold (low gene expression profile pre-DAA therapy), post-hot (high gene expression post-DAA therapy), and post-cold (low gene expression post-DAA therapy).

**Supplementary Figure 4.** The Venn diagram illustrates genes significantly expressed across the pre- and post-DAA therapy subcluster groups compared to the control group. Differentially expressed genes with an adjusted (BY) p-value < 0.01 and a log2-fold change ≥ 0.6 were selected to create the Venn diagram. The post-cold subcluster group was not included because no genes were significantly expressed compared to the control. Genes in red and blue represent those up-regulated or down-regulated, respectively. Abbreviations: Pre-hot (high gene expression profile pre-DAA therapy), pre-cold (low gene expression profile pre-DAA therapy), post-hot (high gene expression post-DAA therapy).

**Supplementary Figure 5. T cell protein expression patterns pre and post-DAA treatment. (A)** Unstained slides were stained with a multiplex T cell panel (CD3+, CD4+, CD8+, CD45RO+, FoxP3+, and DAPI). The Multicomponent TIFF images were then analyzed with Visiopharm to determine the type and number of T cell phenotypes in the portal tracts. The data were used to construct t-SNE plots for individual controls and patients pre- and post-DAA therapy with or without persistent inflammation. Each colored dot represents a unique cellular phenotype, and gray dots represent negative cells for all the markers evaluated. (**B**) These images represent the phenotypic distribution in the portal tract of the multiplex T cell panel (CD3+, CD4+, CD8+, CD45RO+, FoxP3+, and DAPI) from controls and patients pre-DAA, and post-DAA therapy, with or without persistent inflammation. Each colored dot represents a unique cellular phenotype, and gray dots represent negative cells for all the markers evaluated. Abbreviations: Pre-,pre-DAA therapy; Post-,post-DAA therapy; PI, persistent inflammation; NPI, not persistent inflammation.

**Supplementary Figure 6. Box plots comparing the gene expression of CD4, CD8, FoxP3 before and after treatment with DAA**. There was no significant difference in CD4, CD8, and FoxP3 gene expression between patients who were and were not coinfected with HIV before and after treatment. Normal distribution was tested, and an unpaired t-test was used to compare the differences between the groups. *p < 0.05; **p < 0.01; ***p < 0.001. Abbreviations: Ctrl, (controls); Pre-HIV, (Patient with HIV and pre-DAA treatment); Pre-HIV-neg, (Patient without HIV and pre-DAA treatment); Post-HIV, (Patient with HIV and post-DAA treatment); Post-HIV-neg, (Patient without HIV and post-DAA treatment).

**Supplementary Figure 7. Gene expression patterns in patients with HCV are not significantly affected by their HIV status.** To determine whether HIV co-infection was a confounding factor in the observed gene expression differences before and after treatment, we conducted a PCA (**A, B**) and a Fisher’s exact test analysis (**C, D**) in groups of patients with HCV pre-(**A, C;** pre-hot, n = 9; pre-cold, n = 8) and post-(**B, D;** post-hot, n = 3; post-cold, n = 11) treatment compared to HIV status (+/-). Patients co-infected with HIV exhibited overlapping clustering with non-co-infected patients pre-(**A)** and post-(**B**) treatment, and the clustering was influenced by cold or hot status both before and after treatment, as previously demonstrated. Gene expression patterns in patients with HCV pre-(**C**) and post-(**D**) treatment were not significantly influenced by HIV status (Fisher’s test, p > 0.05).

**Table S1.**
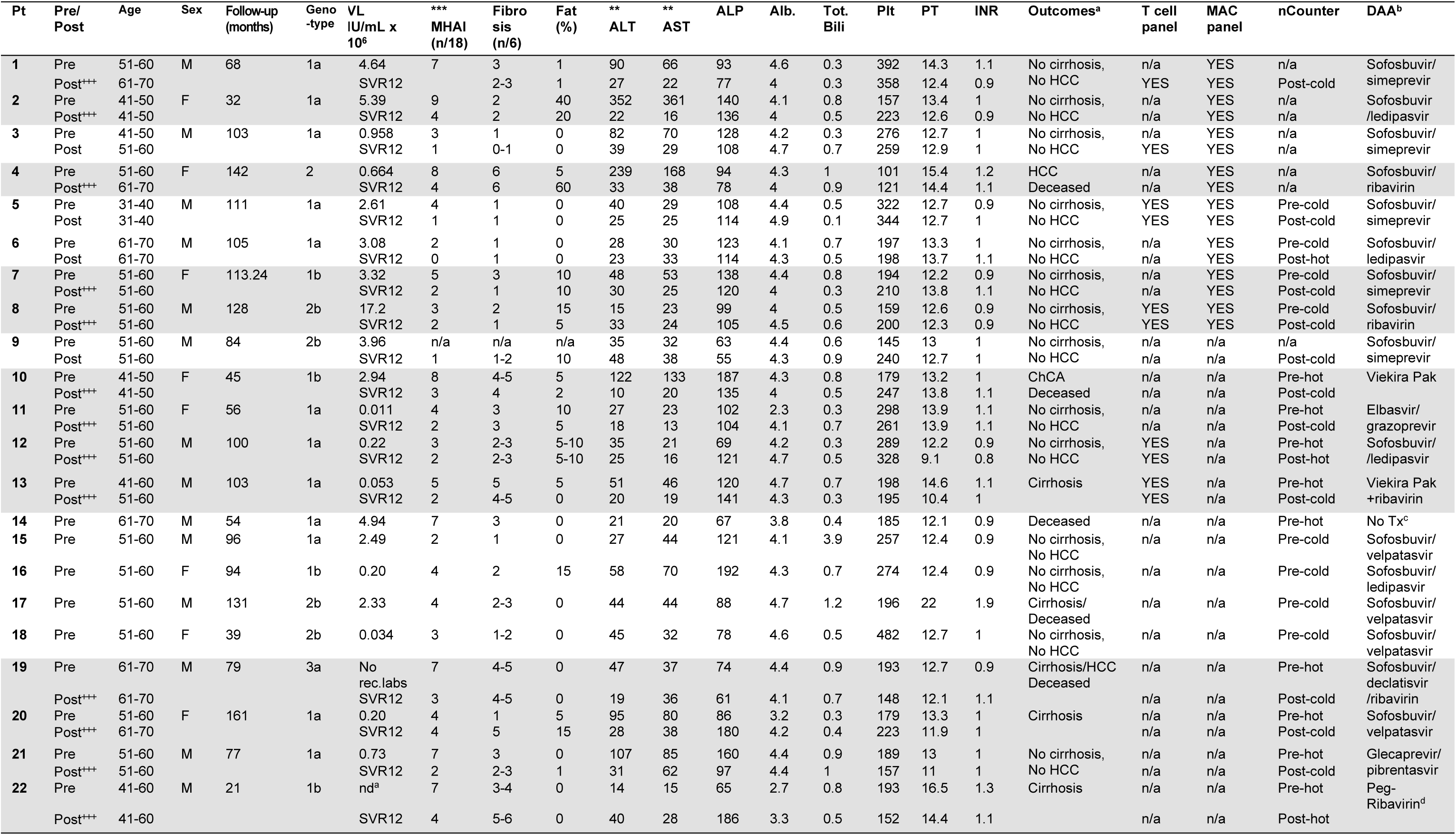
Study Patient Set 1, 2, and 3: Demographic and laboratory data for patients with HCV pre- and post-DAA treatment analyzed with nCounter and multiplexing imaging. Normal range: ALT, 9-51 U/L; AST,13-40 U/L; ALP, 34-122 U/L; Albumin, 3.5-5 g/dL; Bilirubin, 0.1-1.1 mg/dL; Platelets,166-358 x1000/µL; PT, 11-13.5 sec; INR, 1.1. Abbreviations: Alb., albumin; Pt #, patient number; DAA, Direct-actin antivirals; Pre, pre-treatment; Post, post-treatment; Post^+++^, patients with persistent inflammation post-DAA; F, female; M, male; ChCA, cholangiocarcinoma; HCV, hepatitis C virus; HCV gen, HCV genotype; nd, not detected; MHAI, modified hepatic activity index (0-18); No rec.labs, No recent labs; ALT, Alanine transaminase; AST, Aspartate aminotransferase; ALP, Alkaline phosphatase; Plt, Platelets; PT, Prothrombin time; INR, International Normalized Ratio; Tot. bil., total bilirubin; MAC, macrophage. Outcomes: HCC, Hepatocellular carcinoma; n/a, no tissue available; Asterisk on top of MHAI, ALT, and AST represent significant statistical differences between the groups. A normal distribution was tested and an unpaired t-test or a Mann-Whitney (MW) test, as appropriate, with Holm-Sidak correction was used to compare the differences between the groups. *p < 0.05; **p < 0.01; ***p < 0.001; ****p < 0.0001. Superscript a: Patient 4 and 10 succumbed to HCC and ChCA, respectively, while patients 14 and 19 die of cirrhosis-related complications. Patient 13 die of cardiogenic shock. b: All patients completed the duration of the treatment according to the guidelines at the time of treatment except patient 22 and 14. c: Patient did not receive any treatment, d: Treatment discontinue after 5 weeks due to renal complications.

