## Supplementary Tables for "Alterations in the hepatic microenvironment following direct-acting antiviral therapy for chronic hepatitis C"

**Changes in the hepatic microenvironment after direct-acting antiviral therapy for viral hepatitis C.**

**Table of contents**

Table S1……………………………………………………………………………………….…...2

Table S2……………..……………………………………………………………………….…....2

Table S3……………………………………………………………………………………...........3

Table S4……………………………………………………………………………………...........4

Table S5……………………………………………………..………………………………….....5-6

Table S6……………………………………..………………………………………………….....7-10

Table S7…………..……………………………………………………………………………….11

Table S8……………………………………………………………………..………………….....11

Table S9………..……………………………………………………………………………........12-14

Table S10..………………………………………………....…………………………………......15-16

Table S11……………………………………………………………………………………….…..17

Table S12……………..……………………………………………………………………….…...18-19

**Table S1.** Antibodies and optimized multiplex conditions used to identify human macrophage phenotypes and druggable targets in human formalin-fixed, paraffin- embedded liver biopsy tissues.

| **Antibody** | **Vendor/Clone/Isotype** | **Dilution** | **Incubation Time (min)** | **Antigen Retrieval Buffer** | **Opal/**  **Dilution** | **Reaction Position** |
| --- | --- | --- | --- | --- | --- | --- |
| **Macrophage phenotype Panel** | | | | | | |
| **CD68** | Biogenex/KP1/mouse-IgG1κ | RTU* | 60 | AR9 (Akoya) | 520/1:300 | 1 |
| **MAC387** | Dako/Mac387/mouse-IgG1κ | 1:200 | 30 | AR6 (Biogenex) | 690/1:100 | 2 |
| **CD16** | Abcam/EPR16784/rabbit-IgG | 1:500 | 60 | AR6 (Biogenex) | 620/1:600 | 3 |
| **CD14** | Abcam/EPR3653/rabbit-IgG | 1:500 | 45 | AR6 (Biogenex) | 540/1:600 | 4 |
| **CD163** | Leica/10D6/mouse-IgG1 | RTU* | 30 | AR6 (Biogenex) | 650/1:100 | 5 |
| **DAPI** | Akoya | 15µL/mL | 4 | AR6 (Biogenex) | - | 6 |

**Table S2.** Antibodies and optimized multiplex conditions used to identify human T cell phenotypes in human formalin-fixed, paraffin-embedded liver biopsy tissues.

| **Antibody** | **Vendor/Clone/**  **isotype** | **Dilution** | **Incubation Time (min)** | **Antigen Retrieval Buffer** | **Opal/**  **Dilution** | **Reaction Position** |
| --- | --- | --- | --- | --- | --- | --- |
| **T cell phenotype panel** | | | | | | |
| **CD8** | Leica/4B11/IgG2b | 1:4 | 45 | AR9 (Akoya) | 570/1:200 | 1 |
| **CD4** | Leica/4B12/IgG1 | RTU | 60 | AR9 (Akoya) | 520/1:50 | 2 |
| **CD3** | Leica/LN10/IgG1 | RTU | 45 | AR6 (Biogenex) | 650/1:100 | 3 |
| **FoxP3** | CST/D608R/IgG | 1:250 | 30 | AR6 (Biogenex) | 620/1:250 | 4 |
| **CD45RO** | Leica/UCHL1/IgG2a | RTU | 45 | AR6 (Biogenex) | 690/1:100 | 5 |
| **DAPI** | Akoya | 15µL/mL | 5 | AR6 (Biogenex) | - | 6 |

**Table S3-S10: nCounter (NanoString) gene expression results.** Gene expression analysis was performed in patients with HCV pre-DAA treatment (n = 17), post-DAA treatment (n=14) and controls (n = 9) using the PanCancer immune profiling panel, the nCounter sprint profiler and the nSolver advanced analysis module. Log2 FC and the adjusted (p-value<0.05) are shown.

**Table S3.** HCV patients pre-DAA treatment (n = 17) compared to controls (n = 9)

(volcano plot Fig. 2C)

|  | **Log2 fold change** | **BY.p.value (p<0.05)** |
| --- | --- | --- |
| **ISG15** | 3.11 | 2.36E-05 |
| **HLA-B** | 1.61 | 2.36E-05 |
| **STAT2** | 0.778 | 3.28E-05 |
| **HLA-A** | 1.65 | 3.28E-05 |
| **MX1** | 2.58 | 0.000105 |
| **HLA-E** | 0.805 | 0.000389 |
| **IFI27** | 2.95 | 0.000545 |
| **IFI35** | 1.41 | 0.0011 |
| **BST2** | 1.08 | 0.00119 |
| **CASP8** | 0.513 | 0.00202 |
| **HLA-DRB3** | 1.49 | 0.00458 |
| **IFI16** | 1.13 | 0.00458 |
| **IFNAR2** | 0.407 | 0.00458 |
| **PSMB9** | 1.16 | 0.00458 |
| **CD44** | 1.21 | 0.00458 |
| **IFIT1** | 1.68 | 0.00458 |
| **CXCL10** | 3.06 | 0.00468 |
| **LAMP2** | 0.608 | 0.00622 |
| **CXCL9** | 2.33 | 0.00796 |
| **ITGAL** | 0.783 | 0.00894 |
| **HLA-C** | 1.09 | 0.00918 |
| **OAS3** | 1.79 | 0.0148 |
| **C8G** | 0.776 | 0.0148 |
| **STAT5B** | -0.727 | 0.0204 |
| **TNFRSF1B** | 0.688 | 0.0204 |
| **CD24** | 1.84 | 0.0204 |
| **MAP2K1** | -1.04 | 0.0204 |
| **ICAM3** | -0.83 | 0.0204 |
| **CYLD** | 0.594 | 0.0216 |
| **THBS1** | -1.89 | 0.0234 |
| **IL32** | 1.34 | 0.0234 |
| **TAPBP** | 0.947 | 0.028 |
| **RPS6** | -0.676 | 0.0347 |
| **TGFB1** | 0.799 | 0.0406 |
| **CCL19** | 2.05 | 0.0439 |

**Table S4.** HCV patients pre-DAA (n = 17) compared to post-DAA (n = 14) treatment

(volcano plot Fig. 2C)

|  | **Log2 fold change** | **BY.p.value**  **(p<0.05)** |
| --- | --- | --- |
| **ISG15** | 3.01 | 1.32E-05 |
| **MX1** | 2.64 | 3.04E-05 |
| **HLA-B** | 1.4 | 0.000102 |
| **OAS3** | 2.34 | 0.000262 |
| **IFIT1** | 1.73 | 0.00144 |
| **IFI27** | 2.09 | 0.00876 |
| **STAT2** | 0.515 | 0.0124 |
| **HLA-A** | 1.12 | 0.0134 |
| **STAT1** | 1.59 | 0.0203 |
| **CXCL10** | 2.66 | 0.0299 |
| **IFI35** | 1.06 | 0.0372 |

**Table S5.** HCV patients pre-hot (n = 9) compared to pre-cold (n = 8)

(volcano plot Fig. 4A)

|  | **Log2 fold change** | **BY.p.value**  **(p<0.05)** |
| --- | --- | --- |
| **CXCR4** | 2.92 | 0.00223 |
| **FYN** | 1.03 | 0.00257 |
| **BCL2** | 1.53 | 0.00373 |
| **CD74** | 1.41 | 0.00373 |
| **C8B** | -0.711 | 0.00373 |
| **SERPING1** | -0.557 | 0.00436 |
| **CD79B** | 1.55 | 0.00488 |
| **LTB** | 2.34 | 0.00893 |
| **CCL3** | 1.07 | 0.00895 |
| **ITGA4** | 1.4 | 0.0118 |
| **IKBKE** | 1.84 | 0.0165 |
| **IRF7** | 1.06 | 0.0165 |
| **CD24** | 1.56 | 0.0171 |
| **IL13RA1** | -0.453 | 0.0175 |
| **NLRC5** | 1.23 | 0.018 |
| **CSF2RB** | 1.24 | 0.018 |
| **HLA-DMA** | 0.989 | 0.018 |
| **JAK3** | 2.21 | 0.0199 |
| **IL10RA** | 0.932 | 0.0259 |
| **ENTPD1** | 2.14 | 0.0259 |
| **C4BPA** | -0.629 | 0.0259 |
| **TNFAIP3** | 0.935 | 0.0259 |
| **CASP8** | 0.383 | 0.0259 |
| **C1S** | -0.832 | 0.0259 |
| **C3** | -1.12 | 0.0259 |
| **ZAP70** | 1.02 | 0.0259 |
| **IL2RB** | 1.56 | 0.026 |
| **ISG20** | 1.23 | 0.026 |
| **TXNIP** | 0.981 | 0.0292 |
| **IL7R** | 2.38 | 0.0303 |
| **BATF** | 0.925 | 0.0365 |
| **JAK2** | 0.675 | 0.0365 |
| **IL12RB1** | 1.33 | 0.0373 |
| **CD96** | 1.49 | 0.0373 |
| **C8A** | -1.19 | 0.0394 |
| **RUNX3** | 1.94 | 0.0394 |
| **CD8A** | 1.43 | 0.0394 |
| **IRF4** | 1.87 | 0.04 |
| **FN1** | -1.11 | 0.0431 |
| **MAP2K1** | -0.943 | 0.0431 |
| **SBNO2** | 0.586 | 0.0431 |
| **ITGA1** | -0.505 | 0.0431 |
| **LRP1** | -0.288 | 0.0442 |
| **IKBKB** | 0.546 | 0.0453 |
| **CXCL9** | 1.94 | 0.0469 |
| **LAIR2** | 1.08 | 0.0472 |
| **LILRB2** | 0.829 | 0.0483 |

**Table S6.** HCV patients pre-hot (n = 9) compared to controls (n = 9)

(volcano plot Fig. 4C, Fig. S4)

|  | **Log2 fold change** | **BY.p.value** |
| --- | --- | --- |
| **ISG20** | 2.55 | 2.99E-07 |
| **HLA-B** | 1.9 | 2.17E-06 |
| **CD8A** | 2.79 | 4.50E-06 |
| **C8B** | -0.967 | 4.50E-06 |
| **BCL2** | 1.85 | 4.50E-06 |
| **ISG15** | 3.44 | 4.50E-06 |
| **FYN** | 1.17 | 4.50E-06 |
| **CASP8** | 0.677 | 4.50E-06 |
| **TNFAIP3** | 1.49 | 4.50E-06 |
| **HLA-A** | 1.89 | 5.60E-06 |
| **STAT2** | 0.861 | 8.54E-06 |
| **CSF2RB** | 1.72 | 1.27E-05 |
| **ZAP70** | 1.8 | 1.30E-05 |
| **IKBKB** | 0.912 | 1.30E-05 |
| **HLA-E** | 0.962 | 1.42E-05 |
| **LILRB2** | 1.38 | 1.42E-05 |
| **CD44** | 1.54 | 1.79E-05 |
| **HLA-DRB3** | 1.81 | 1.79E-05 |
| **TAP1** | 1.5 | 1.79E-05 |
| **IRF4** | 2.95 | 1.97E-05 |
| **IFI16** | 1.47 | 3.03E-05 |
| **CD24** | 2.38 | 3.94E-05 |
| **MAP2K1** | -1.56 | 3.95E-05 |
| **MX1** | 2.83 | 3.95E-05 |
| **BATF** | 1.33 | 3.95E-05 |
| **C3** | -1.54 | 3.95E-05 |
| **LTB** | 2.66 | 6.78E-05 |
| **IRF7** | 1.45 | 6.78E-05 |
| **CXCL9** | 2.95 | 6.78E-05 |
| **BST2** | 1.29 | 8.00E-05 |
| **TAP2** | 1.36 | 8.26E-05 |
| **CD79B** | 1.56 | 8.56E-05 |
| **TNFRSF1B** | 0.926 | 9.84E-05 |
| **CCL5** | 1.61 | 0.000121 |
| **ICAM3** | -1.18 | 0.000145 |
| **TGFB1** | 1.04 | 0.000167 |
| **IFNAR2** | 0.498 | 0.000174 |
| **NFKB2** | 1.33 | 0.000174 |
| **CXCL10** | 3.55 | 0.000224 |
| **ITGA4** | 1.46 | 0.000234 |
| **ITGAL** | 0.99 | 0.000262 |
| **ITGA1** | -0.627 | 0.000268 |
| **HMGB1** | -1.19 | 0.000268 |
| **IFI27** | 3.15 | 0.000277 |
| **IFI35** | 1.55 | 0.000335 |
| **ATG5** | -0.981 | 0.000335 |
| **ITCH** | -0.731 | 0.00034 |
| **CD81** | -0.829 | 0.00034 |
| **FN1** | -1.38 | 0.00034 |
| **CD3E** | 1.92 | 0.000348 |
| **NLRC5** | 1.43 | 0.000348 |
| **JAK2** | 0.829 | 0.000348 |
| **IKBKE** | 1.98 | 0.000375 |
| **C4BPA** | -0.762 | 0.000387 |
| **LAMP2** | 0.742 | 0.000387 |
| **REPS1** | -1.18 | 0.00045 |
| **ATG7** | 0.769 | 0.00045 |
| **CXCL11** | 2.19 | 0.000497 |
| **PSMB9** | 1.35 | 0.00055 |
| **STAT5B** | -0.899 | 0.000573 |
| **ATF1** | -0.687 | 0.000604 |
| **SIGLEC1** | 0.826 | 0.000612 |
| **CCL3** | 0.992 | 0.000612 |
| **CD84** | 1.15 | 0.000659 |
| **HLA-DMA** | 1.01 | 0.000661 |
| **LRP1** | -0.38 | 0.000661 |
| **JAK3** | 2.24 | 0.000729 |
| **IFNAR1** | -0.655 | 0.000763 |
| **CD96** | 1.67 | 0.000888 |
| **CD74** | 1.37 | 0.0011 |
| **IFIH1** | 1.32 | 0.00113 |
| **AMBP** | -1.03 | 0.0012 |
| **CCL19** | 2.67 | 0.00125 |
| **CD164** | -0.874 | 0.00125 |
| **CXCR4** | 2.08 | 0.00125 |
| **IL12RB1** | 1.48 | 0.00125 |
| **RORC** | -0.862 | 0.00134 |
| **CCRL2** | -0.791 | 0.00139 |
| **CD27** | 2.79 | 0.00196 |
| **CD99** | 0.726 | 0.00227 |
| **TNFSF13** | 1.7 | 0.00232 |
| **IL13RA1** | -0.415 | 0.00238 |
| **C1R** | -0.673 | 0.00238 |
| **TRAF3** | 0.791 | 0.00251 |
| **OAS3** | 2.03 | 0.00251 |
| **HLA-C** | 1.18 | 0.0026 |
| **RUNX3** | 1.99 | 0.00303 |
| **IFIT1** | 1.73 | 0.00331 |
| **KIT** | -1.52 | 0.00352 |
| **CD9** | -1.16 | 0.00372 |
| **IL18** | 1.35 | 0.00398 |
| **IGF1R** | 0.837 | 0.0044 |
| **IL13RA2** | -1.88 | 0.0044 |
| **TFEB** | 0.906 | 0.00458 |
| **IL32** | 1.55 | 0.00459 |
| **CD5** | 1.56 | 0.00487 |
| **PIK3CG** | 1.57 | 0.00487 |
| **SBNO2** | 0.65 | 0.00487 |
| **CD79A** | 1.79 | 0.00506 |
| **CD46** | -0.645 | 0.00508 |
| **CTSH** | -1.05 | 0.00508 |
| **C8G** | 0.832 | 0.00574 |
| **MAPKAPK2** | 0.561 | 0.00574 |
| **CLEC4A** | 0.528 | 0.00574 |
| **IRF3** | 1.06 | 0.00574 |
| **CYLD** | 0.668 | 0.00604 |
| **STAT1** | 1.58 | 0.00604 |
| **C6** | -0.71 | 0.00639 |
| **TICAM1** | 0.603 | 0.00655 |
| **C1S** | -0.756 | 0.0069 |
| **ST6GAL1** | -0.798 | 0.00749 |
| **PRKCD** | 0.722 | 0.00763 |
| **IL10RA** | 0.772 | 0.0078 |
| **C8A** | -1.08 | 0.00796 |
| **NLRP3** | 1.19 | 0.0087 |
| **CTSL** | -0.935 | 0.00894 |
| **SAA1** | 2.86 | 0.00901 |
| **PDGFC** | -0.653 | 0.0092 |
| **LAIR2** | 1.18 | 0.00926 |
| **STAT6** | 0.738 | 0.00932 |
| **TXNIP** | 0.882 | 0.00934 |
| **IKBKG** | 0.771 | 0.00934 |
| **CCL21** | 2.01 | 0.0105 |
| **CREBBP** | -0.725 | 0.011 |
| **TNFSF4** | 0.864 | 0.0113 |
| **CEBPB** | -0.855 | 0.0113 |
| **THBS1** | -2.03 | 0.0117 |
| **TAPBP** | 0.958 | 0.0132 |
| **ATF2** | -0.363 | 0.0133 |
| **ENTPD1** | 1.67 | 0.0158 |
| **NFATC3** | -0.592 | 0.0162 |
| **NFATC2** | 0.651 | 0.0184 |
| **CD3D** | 1.82 | 0.0187 |
| **HLA-G** | 1.4 | 0.0187 |
| **CCL17** | -1.1 | 0.0187 |
| **CFD** | 1.04 | 0.0192 |
| **TIGIT** | 1.29 | 0.0199 |
| **CCL14** | -0.854 | 0.0204 |
| **TLR2** | 0.788 | 0.0206 |
| **PTPRC** | 0.798 | 0.0211 |
| **GZMK** | 1.53 | 0.0218 |
| **CEACAM1** | 1.07 | 0.0218 |
| **AKT3** | 0.53 | 0.022 |
| **YTHDF2** | -0.563 | 0.0226 |
| **SMAD2** | -0.821 | 0.0238 |
| **CCL3L1** | 1.16 | 0.0286 |
| **HLA-DOB** | 1.3 | 0.0305 |
| **SERPING1** | -0.363 | 0.0313 |
| **TARP** | 1.26 | 0.032 |
| **TANK** | -0.468 | 0.0341 |
| **IL18R1** | 0.651 | 0.0369 |
| **MAP4K2** | 0.601 | 0.0376 |
| **CDKN1A** | 1.11 | 0.0382 |
| **IRAK4** | 0.563 | 0.0384 |
| **IL17RB** | 0.933 | 0.0397 |
| **IRF1** | 0.804 | 0.0417 |
| **ALCAM** | -0.605 | 0.0423 |
| **DDX58** | 0.845 | 0.0427 |
| **MAPK1** | -0.57 | 0.0446 |
| **C5** | -0.424 | 0.0446 |
| **A2M** | 0.864 | 0.0473 |
| **ITGB3** | 0.7 | 0.0475 |

**Table S7.** HCV patients pre- cold (n = 8) compared to controls (n = 9)

(volcano plot Fig. 4C, Fig. S4)

|  | **Log2 fold change** | **BY.p.value** |
| --- | --- | --- |
| **ISG15** | 2.6 | 0.0069 |
| **STAT2** | 0.679 | 0.0069 |
| **HLA-B** | 1.2 | 0.016 |
| **HLA-A** | 1.32 | 0.016 |
| **MX1** | 2.23 | 0.018 |
| **IFI27** | 2.68 | 0.0301 |
| **ISG20** | 1.32 | 0.0301 |

**Table S8.** HCV patients post-hot (n = 3) compared to post-cold (n = 11)

(volcano plot Fig. 4B)

|  | **Log2 fold change** | **BY.p.value** |
| --- | --- | --- |
| **SERPING1** | -0.901 | 0.00138 |
| **C8B** | -1.03 | 0.00208 |
| **ITCH** | -1.16 | 0.00208 |
| **KIT** | -4.22 | 0.00345 |
| **NFATC3** | -1.24 | 0.0136 |
| **ATG5** | -1.43 | 0.0166 |
| **CD164** | -1.27 | 0.0166 |
| **C1S** | -1.25 | 0.0166 |
| **CD46** | -1.09 | 0.0166 |
| **C3** | -1.66 | 0.0166 |
| **CD24** | 2.11 | 0.0166 |
| **AMBP** | -1.48 | 0.0166 |
| **BMI1** | -0.913 | 0.0168 |
| **CD40** | -1 | 0.0168 |
| **CD81** | -1.09 | 0.0197 |
| **ITGA1** | -0.796 | 0.02 |
| **IRAK4** | -1.55 | 0.02 |
| **FN1** | -1.72 | 0.02 |
| **YTHDF2** | -1.05 | 0.0212 |
| **PDGFC** | -1.11 | 0.0215 |
| **CFB** | -1.06 | 0.0226 |
| **MRC1** | -1.13 | 0.0234 |
| **RORC** | -1.16 | 0.0318 |
| **CTSH** | -1.58 | 0.0377 |
| **SMAD2** | -1.43 | 0.0377 |
| **IL13RA1** | -0.533 | 0.0436 |
| **TGFB1** | 1.03 | 0.0446 |

**Table S9.** HCV patients post-hot (n = 3) compared to controls (n = 9)

(volcano plot Fig. 4C, Fig. S4)

|  | **Log2 fold change** | **BY.p.value** |
| --- | --- | --- |
| **ITCH** | -1.42 | 5.40E-05 |
| **C8B** | -1.27 | 7.99E-05 |
| **ATG5** | -1.82 | 0.000134 |
| **KIT** | -4.41 | 0.000134 |
| **SERPING1** | -0.962 | 0.000134 |
| **TGFB1** | 1.58 | 0.000223 |
| **HLA-DRB3** | 2.32 | 0.000294 |
| **FN1** | -2.17 | 0.000313 |
| **C3** | -2.07 | 0.000313 |
| **CD81** | -1.29 | 0.000313 |
| **NFATC3** | -1.37 | 0.000365 |
| **RORC** | -1.49 | 0.000419 |
| **CD164** | -1.49 | 0.000419 |
| **ITGA1** | -0.93 | 0.000419 |
| **BMI1** | -1.05 | 0.000525 |
| **C1R** | -1.16 | 0.00068 |
| **IL6ST** | -1.08 | 0.000836 |
| **IGF1R** | 1.45 | 0.000864 |
| **CD24** | 2.83 | 0.000864 |
| **AMBP** | -1.61 | 0.000907 |
| **CD44** | 1.71 | 0.00101 |
| **STAT5B** | -1.31 | 0.00148 |
| **CTSH** | -1.82 | 0.00155 |
| **CD46** | -1.1 | 0.00173 |
| **C1S** | -1.31 | 0.00201 |
| **SMAD2** | -1.7 | 0.00222 |
| **IRAK4** | -1.62 | 0.00246 |
| **STAT6** | 1.27 | 0.00289 |
| **YTHDF2** | -1.11 | 0.00289 |
| **IFITM1** | -2.47 | 0.00289 |
| **MAPK1** | -1.21 | 0.00289 |
| **CTSL** | -1.61 | 0.00289 |
| **REPS1** | -1.54 | 0.00296 |
| **ATF1** | -0.936 | 0.00335 |
| **TICAM2** | -2.62 | 0.00335 |
| **EP300** | -0.908 | 0.00393 |
| **CD99** | 1.04 | 0.00393 |
| **PDGFC** | -1.13 | 0.00393 |
| **ATG7** | 0.938 | 0.00393 |
| **MRC1** | -1.13 | 0.00606 |
| **TP53** | -0.97 | 0.00695 |
| **ATG10** | -2.17 | 0.00925 |
| **HMGB1** | -1.34 | 0.00927 |
| **IKBKB** | 0.791 | 0.00954 |
| **RIPK2** | -2.61 | 0.00977 |
| **HLA-DRA** | -1.94 | 0.00999 |
| **CD63** | 1.28 | 0.0109 |
| **ST6GAL1** | -1.17 | 0.0109 |
| **ATF2** | -0.576 | 0.0121 |
| **TFRC** | -1.35 | 0.0124 |
| **C4BPA** | -0.823 | 0.0168 |
| **TLR1** | -1.67 | 0.0169 |
| **BCL2** | 1.4 | 0.017 |
| **CCL14** | -1.34 | 0.0176 |
| **CD58** | -1.42 | 0.0187 |
| **TAPBP** | 1.41 | 0.0188 |
| **CREB1** | -1.09 | 0.0188 |
| **TIGIT** | -3.76 | 0.0188 |
| **BATF** | 1.13 | 0.0203 |
| **TNFSF13** | 1.98 | 0.0219 |
| **AKT3** | 0.803 | 0.0219 |
| **LY86** | -2.74 | 0.0224 |
| **CASP3** | -1.7 | 0.0227 |
| **C5** | -0.706 | 0.0231 |
| **LY96** | -1.66 | 0.0231 |
| **CKLF** | -1.32 | 0.0237 |
| **IFITM2** | -0.881 | 0.0249 |
| **IL12RB1** | 1.6 | 0.0261 |
| **CCL3** | 1.01 | 0.0264 |
| **CD40** | -0.938 | 0.0311 |
| **IL13RA2** | -2.38 | 0.032 |
| **FCGR2B** | -1.15 | 0.032 |
| **MAP2K2** | 0.744 | 0.0323 |
| **CCR2** | -4.22 | 0.0324 |
| **IL13RA1** | -0.47 | 0.0363 |
| **APP** | -0.749 | 0.0368 |
| **TNFRSF1B** | 0.793 | 0.0374 |
| **CLEC4A** | 0.618 | 0.0386 |
| **CYFIP2** | -1.67 | 0.0394 |
| **PSEN1** | -1.47 | 0.0421 |
| **PLA2G6** | 0.869 | 0.0421 |
| **FCGR3A** | -1.39 | 0.0421 |
| **C4B** | 1.2 | 0.0452 |
| **CX3CL1** | -1.26 | 0.0468 |
| **TIRAP** | 0.615 | 0.0487 |
| **C8G** | 0.98 | 0.0487 |
| **ATG16L1** | -0.881 | 0.0487 |
| **CD53** | -2.1 | 0.0487 |

**Table S10.** HCV patients pre-hot (n = 9) compared to post-cold (n = 11)

|  | **Log2 fold change** | **BY.p.value** |
| --- | --- | --- |
| **HLA-B** | 1.69 | 3.78E-05 |
| **IRF7** | 1.62 | 3.78E-05 |
| **ISG15** | 3.29 | 3.78E-05 |
| **ISG20** | 1.95 | 9.12E-05 |
| **MX1** | 2.92 | 0.000105 |
| **C8B** | -0.724 | 0.000706 |
| **BCL2** | 1.49 | 0.000706 |
| **CD74** | 1.4 | 0.000706 |
| **FYN** | 0.929 | 0.000706 |
| **OAS3** | 2.41 | 0.00111 |
| **CASP8** | 0.477 | 0.00113 |
| **TAP1** | 1.2 | 0.00159 |
| **CXCR4** | 2.32 | 0.00159 |
| **LRP1** | -0.38 | 0.00175 |
| **STAT2** | 0.66 | 0.00193 |
| **CD24** | 1.66 | 0.00231 |
| **BATF** | 1.13 | 0.00244 |
| **IL13RA1** | -0.471 | 0.0029 |
| **NLRC5** | 1.28 | 0.00314 |
| **HLA-C** | 0.996 | 0.00314 |
| **DDX58** | 1.36 | 0.00314 |
| **CD79B** | 1.33 | 0.00368 |
| **IL10RA** | 0.994 | 0.00434 |
| **HLA-E** | 0.705 | 0.00438 |
| **HLA-A** | 1.33 | 0.00443 |
| **C4BPA** | -0.665 | 0.00475 |
| **IKBKB** | 0.638 | 0.00523 |
| **IKBKE** | 1.69 | 0.00617 |
| **ZAP70** | 1.05 | 0.00617 |
| **IFIT1** | 1.71 | 0.00678 |
| **C3** | -1.14 | 0.00692 |
| **TNFAIP3** | 0.943 | 0.00692 |
| **JAK2** | 0.712 | 0.00785 |
| **LTB** | 1.91 | 0.00946 |
| **HLA-DMA** | 0.906 | 0.00955 |
| **CXCL9** | 2.11 | 0.00959 |
| **TXNIP** | 0.976 | 0.00959 |
| **CXCL10** | 2.83 | 0.00959 |
| **STAT1** | 1.76 | 0.00959 |
| **IRF3** | 1.15 | 0.00959 |
| **CXCL11** | 1.95 | 0.0105 |
| **IFI27** | 2.2 | 0.0105 |
| **TAP2** | 0.993 | 0.0113 |
| **BST2** | 0.828 | 0.0116 |
| **ITGA4** | 1.11 | 0.0142 |
| **C8A** | -1.18 | 0.0142 |
| **SIGLEC1** | 0.647 | 0.0142 |
| **IL12RB1** | 1.29 | 0.0144 |
| **SELL** | 1.75 | 0.0167 |
| **AMBP** | -0.896 | 0.0186 |
| **CCL3** | 0.777 | 0.0218 |
| **ITGA1** | -0.487 | 0.0224 |
| **IFI35** | 1.2 | 0.0224 |
| **IFNAR1** | -0.544 | 0.0226 |
| **TNFRSF1B** | 0.611 | 0.0226 |
| **CD84** | 0.944 | 0.0226 |
| **CD46** | -0.626 | 0.0233 |
| **TANK** | -0.529 | 0.0233 |
| **CCRL2** | -0.635 | 0.0248 |
| **IFIH1** | 1.1 | 0.0248 |
| **CD8A** | 1.3 | 0.025 |
| **ALCAM** | -0.737 | 0.0251 |
| **PDGFC** | -0.647 | 0.029 |
| **C1S** | -0.69 | 0.0319 |
| **IRAK4** | 0.644 | 0.0356 |
| **CD81** | -0.624 | 0.0356 |
| **CD96** | 1.27 | 0.0387 |
| **CD3E** | 1.26 | 0.0414 |
| **HMGB1** | -0.811 | 0.0419 |
| **MAP2K1** | -0.828 | 0.0421 |
| **CD5** | 1.43 | 0.0459 |

**Table S11.** HCV patients' post-cold (n = 11) compared to controls (n = 9)

|  | **Log2 fold change** | **BY.p.value** |
| --- | --- | --- |
| **A2M** | 1.17 | 0.157 |
| **ATF1** | -0.543 | 0.175 |
| **ICAM3** | -0.815 | 0.175 |
| **CD8A** | 1.5 | 0.175 |
| **IRF4** | 1.71 | 0.193 |
| **CSF2RB** | 0.908 | 0.23 |
| **NLRP3** | 1.08 | 0.23 |
| **THBS1** | -1.89 | 0.23 |
| **TNFSF4** | 0.797 | 0.253 |
| **LILRB2** | 0.684 | 0.402 |
| **CCL14** | -0.775 | 0.402 |
| **CD44** | 0.744 | 0.402 |
| **STAT5B** | -0.559 | 0.402 |
| **IFI16** | 0.736 | 0.402 |
| **ITGAL** | 0.572 | 0.414 |
| **GTF3C1** | 0.57 | 0.437 |
| **IL18** | 0.963 | 0.437 |
| **C8G** | 0.612 | 0.437 |
| **CXCL2** | -0.999 | 0.437 |
| **HLA-DRB3** | 0.828 | 0.437 |
| **MAP2K1** | -0.733 | 0.49 |
| **TGFB1** | 0.544 | 0.495 |

**Table S12**. Additional clinical information including HIV co-infection status and CD4 counts

| **Pt #** | **Pre/Post** | **HIV** | **HIV log copies/ml** | **CD4**  **(31-60%)** | **CD4 Absolute (410-1,590/cmm)** | **Ab** | **BMI**  **(Kg/m^2^)** | **Duration b/w Pre/Post Bx (months)** | **Duration SVR &**  **Post-Tx Biopsy (months)** | **Duration Dx & Outcome (months)** |
| --- | --- | --- | --- | --- | --- | --- | --- | --- | --- | --- |
| 1 | Pre | - | n/a | n/a | n/a | nd | 25 | 54.8 | 14.2 | 156 |
|  | Post | - | n/a | n/a | n/a |  |  |  |  |  |
| 2 | Pre | **+** | 2471 | 20 | 402 | nd | 45 | 16.6 | 9.4 | 313 |
|  | Post |  | nd | 16 | 526 |  | 46.3 |  |  |  |
| 3 | Pre | **+** | <75 | 14% | 411 | nd | 26.8 | 70.2 | 17.5 | 211 |
|  | Post |  | nd | 16% | 555 |  | 28.7 |  |  |  |
| 4 | Pre | **-** | n/a | n/a | n/a | Pos**^**^** | 32.1 | 85.8 | 28.4 | 142 |
|  | Post | **-** | n/a | n/a | n/a |  |  |  |  |  |
| 5 | Pre | **+** | <75 | 18% | 411 | nd | 21 | 20.8 | 19.2 | 141 |
|  | Post |  | nd | 21 | 409 |  | 23.2 |  |  |  |
| 6 | Pre | **+** | nd | 18% | 375 | nd | 20.7 | 15.3 | 6 | 127 |
|  | Post |  | nd | 27 | 670 |  | 20.6 |  |  |  |
| 7 | Pre | **+** | nd | 52 | 1,034 | nd | 33.6 | 26.8 | 24.1 | 158 |
|  | Post |  | nd | 53 | 1,488 |  | 35.2 |  |  |  |
| 8 | Pre | **+** | 8,758 | 18 | 374 | nd | 23.4 | 24.6 | 20.1 | 128 |
|  | Post |  | nd | 28% | 667 |  | 25.5 |  |  |  |
| 9 | Pre | **+** | <40 | 32 | 446 | nd | 25.9 | ND | 26 | 159 |
|  | Post |  | 66 | 21 | 444 |  | 23.4 |  |  |  |
| 10 | Pre | **+** | nd | 23% | 618 | Pos***** | 28.2 | 21.1 | 15.3 | 176 |
|  | Post |  | nd | 22% | 795 |  | 29 |  |  |  |
| 11 | Pre | **+** | <40 | 20% | 403% | nd | 34.4 | 24 | 86 | 188 |
|  |  |  | nd | 24 | 571 |  | 34.7 |  |  |  |
| 12 | Pre | **+** | nd | 46% | 1,270 | nd | 33.1 | 34.1 | 26 | 242 |
|  |  |  | nd | 51 | 491 |  | 35.9 |  |  |  |
| 13 | Pre | **+** | <40 | 12% | 207 | nd | 25.15 | 45 | 31.21 | 204 |
|  | Post |  | <40 | 19% | 574 |  | 24.48 |  |  |  |
| 14 | Pre | **-** | n/a |  |  | nd | 24.21 | n/a | n/a | 59 |
| 15 | Pre | **+** | nd | 33% | 509 | nd | 24.21 | n/a | n/a | 200.28 |
| 16 | Pre | **-** | n/a | n/a | n/a | nd | 26 | n/a | n/a | 94 |
| 17 | Pre | **-** | n/a | n/a | n/a | nd | 21.14 | n/a | n/a | 131.02 |
| 18 | Pre | **-** | n/a | n/a | n/a | nd | 27.14 | n/a | n/a | 39.2 |
| 19 | Pre | **-** | n/a | n/a | n/a | nd | 27.69 | 68.26 | 62 | 119.19 |
|  | Post | **-** | n/a | n/a | n/a |  | 28.11 |  |  |  |
| 20 | Pre | **-** | n/a | n/a | n/a | nd | 39.36 | 125.26 | 17.13 | 225.22 |
|  | Post | **-** | n/a | n/a | n/a |  | 40.05 |  |  |  |
| 21 | Pre | **-** | n/a | n/a | n/a | ANA | 28.96 | 57 | 9 | 508.25 |
|  | Post | **-** | n/a | n/a | n/a |  | 31.6 |  |  |  |
| 22 | Pre | **-** | n/a | n/a | n/a | nd | 22.29 | 7 | 5.19 | 60 |
|  | Post | **-** | n/a | n/a | n/a | nd |  |  |  |  |

**Abbreviations**: Pt#: Patient numbers (see Table 1 for additional data), Pre/Post: Pre-treatment liver biopsy or Post-treatment liver biopsy; HIV: Human Immunodeficiency Virus; Ab: Antibodies; BMI: body mass index; SVR: Sustained Virologic Response, Bx: Biopsy, Post-Tx: Post-treatment, nd: not detected; n/a: not applicable; Dx: Diagnosis, Duration b/w pre/post bx: Duration between pre and post liver biopsies (in months); Duration of SVR & Post-Tx Biopsy: Duration to achieve SVR and liver biopsy post-treatment (in months), Duration Dx & outcome: Duration between the patient first positive HCV test and latest follow up clinical visit. Pos^*^: IgG 22 (6.18.14), ASM pos 1:20, Pos^**:^ IgG actin (2 low titers), biopsy negative for autoimmune hepatitis.
