## Supplementary material for "Alterations in the hepatic microenvironment following direct-acting antiviral therapy for chronic hepatitis C": Suppl figures

### Slide 1
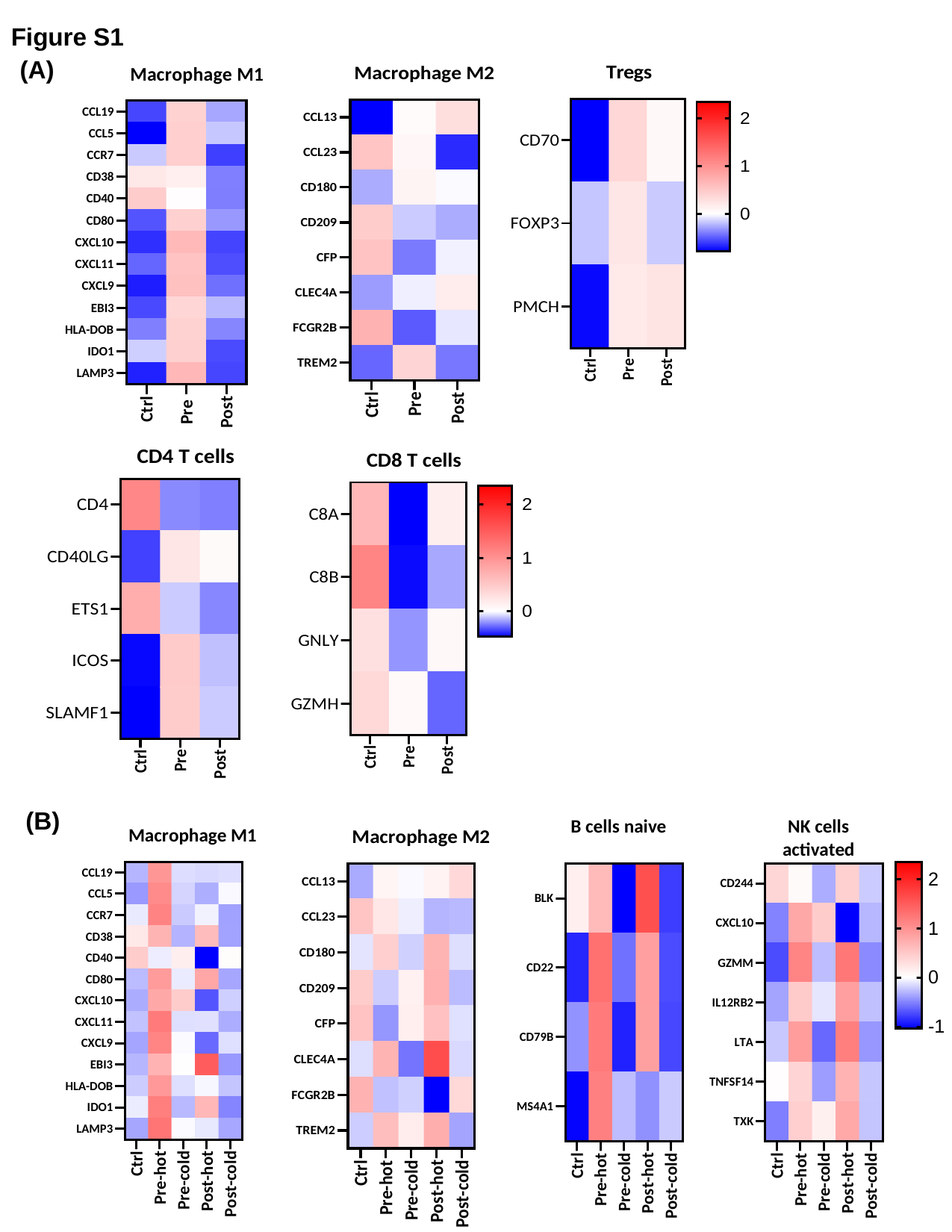

Figure S1
(A)
(B)

### Slide 2
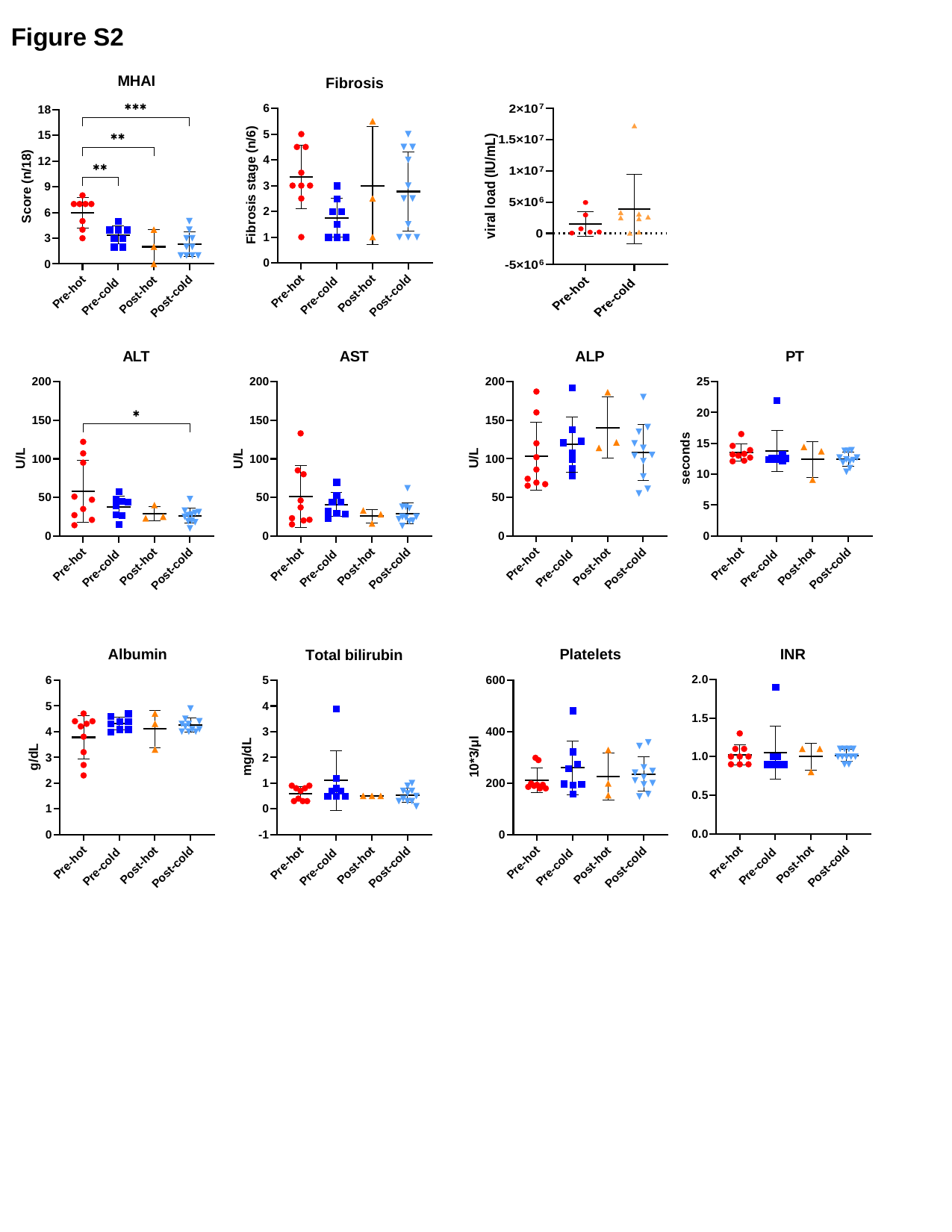

Figure S2

### Slide 3
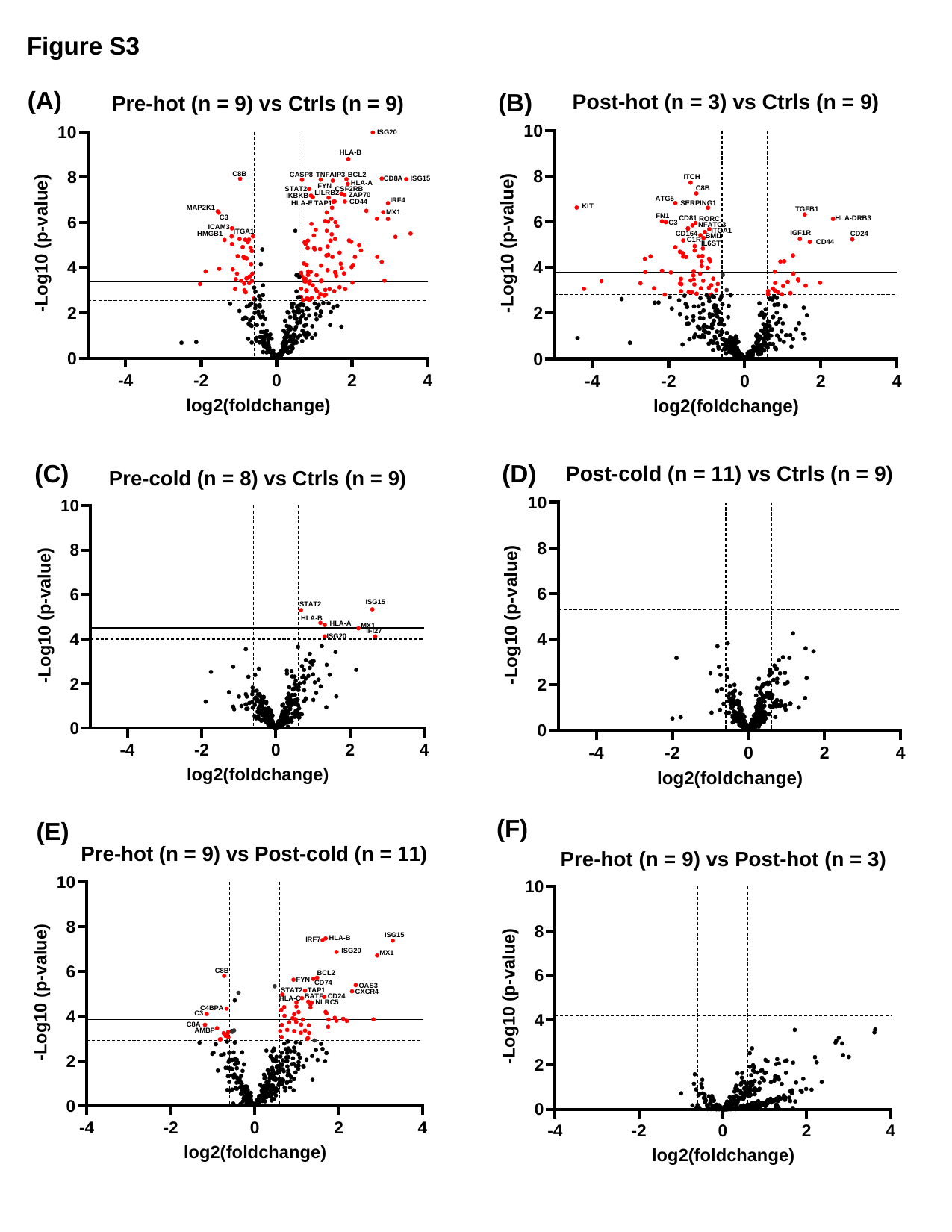

Figure S3
(A)
(B)
(C)
(D)
(F)
(E)

### Slide 4
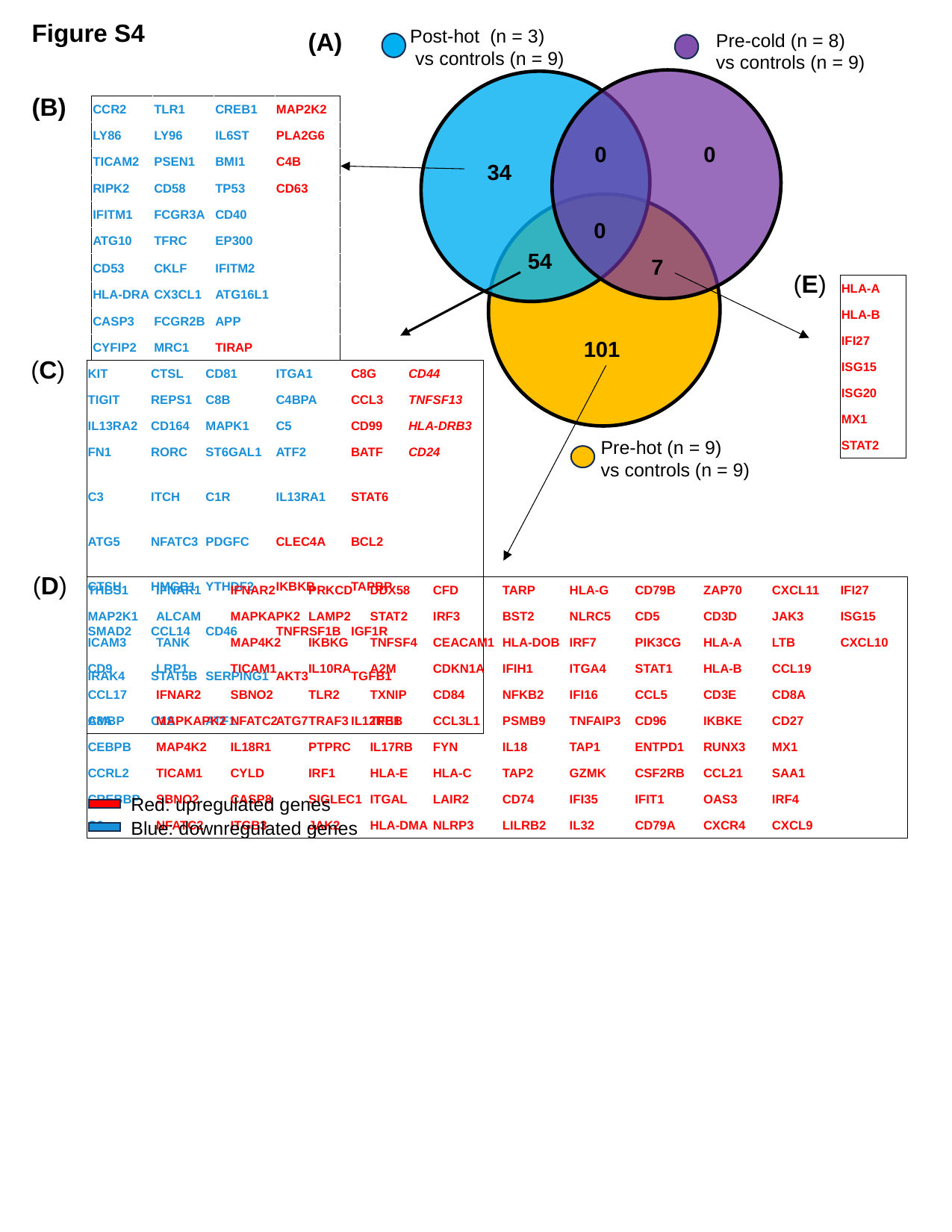

Figure S4
Post-hot (n = 3)
 vs controls (n = 9)
(A)
Pre-cold (n = 8)
vs controls (n = 9)
(B)
| CCR2 | TLR1 | CREB1 | MAP2K2 |
| --- | --- | --- | --- |
| LY86 | LY96 | IL6ST | PLA2G6 |
| TICAM2 | PSEN1 | BMI1 | C4B |
| RIPK2 | CD58 | TP53 | CD63 |
| IFITM1 | FCGR3A | CD40 | |
| ATG10 | TFRC | EP300 | |
| CD53 | CKLF | IFITM2 | |
| HLA-DRA | CX3CL1 | ATG16L1 | |
| CASP3 | FCGR2B | APP | |
| CYFIP2 | MRC1 | TIRAP | |
0
0
34
0
54
7
(E)
| HLA-A |
| --- |
| HLA-B |
| IFI27 |
| ISG15 |
| ISG20 |
| MX1 |
| STAT2 |
101
(C)
| KIT | CTSL | CD81 | ITGA1 | C8G | CD44 |
| --- | --- | --- | --- | --- | --- |
| TIGIT | REPS1 | C8B | C4BPA | CCL3 | TNFSF13 |
| IL13RA2 | CD164 | MAPK1 | C5 | CD99 | HLA-DRB3 |
| FN1 | RORC | ST6GAL1 | ATF2 | BATF | CD24 |
| C3 | ITCH | C1R | IL13RA1 | STAT6 | |
| ATG5 | NFATC3 | PDGFC | CLEC4A | BCL2 | |
| CTSH | HMGB1 | YTHDF2 | IKBKB | TAPBP | |
| SMAD2 | CCL14 | CD46 | TNFRSF1B | IGF1R | |
| IRAK4 | STAT5B | SERPING1 | AKT3 | TGFB1 | |
| AMBP | C1S | ATF1 | ATG7 | IL12RB1 | |
Pre-hot (n = 9)
vs controls (n = 9)
(D)
| THBS1 | IFNAR1 | IFNAR2 | PRKCD | DDX58 | CFD | TARP | HLA-G | CD79B | ZAP70 | CXCL11 | IFI27 |
| --- | --- | --- | --- | --- | --- | --- | --- | --- | --- | --- | --- |
| MAP2K1 | ALCAM | MAPKAPK2 | LAMP2 | STAT2 | IRF3 | BST2 | NLRC5 | CD5 | CD3D | JAK3 | ISG15 |
| ICAM3 | TANK | MAP4K2 | IKBKG | TNFSF4 | CEACAM1 | HLA-DOB | IRF7 | PIK3CG | HLA-A | LTB | CXCL10 |
| CD9 | LRP1 | TICAM1 | IL10RA | A2M | CDKN1A | IFIH1 | ITGA4 | STAT1 | HLA-B | CCL19 | |
| CCL17 | IFNAR2 | SBNO2 | TLR2 | TXNIP | CD84 | NFKB2 | IFI16 | CCL5 | CD3E | CD8A | |
| C8A | MAPKAPK2 | NFATC2 | TRAF3 | TFEB | CCL3L1 | PSMB9 | TNFAIP3 | CD96 | IKBKE | CD27 | |
| CEBPB | MAP4K2 | IL18R1 | PTPRC | IL17RB | FYN | IL18 | TAP1 | ENTPD1 | RUNX3 | MX1 | |
| CCRL2 | TICAM1 | CYLD | IRF1 | HLA-E | HLA-C | TAP2 | GZMK | CSF2RB | CCL21 | SAA1 | |
| CREBBP | SBNO2 | CASP8 | SIGLEC1 | ITGAL | LAIR2 | CD74 | IFI35 | IFIT1 | OAS3 | IRF4 | |
| C6 | NFATC2 | ITGB3 | JAK2 | HLA-DMA | NLRP3 | LILRB2 | IL32 | CD79A | CXCR4 | CXCL9 | |
Red: upregulated genes
Blue: downregulated genes

### Slide 5
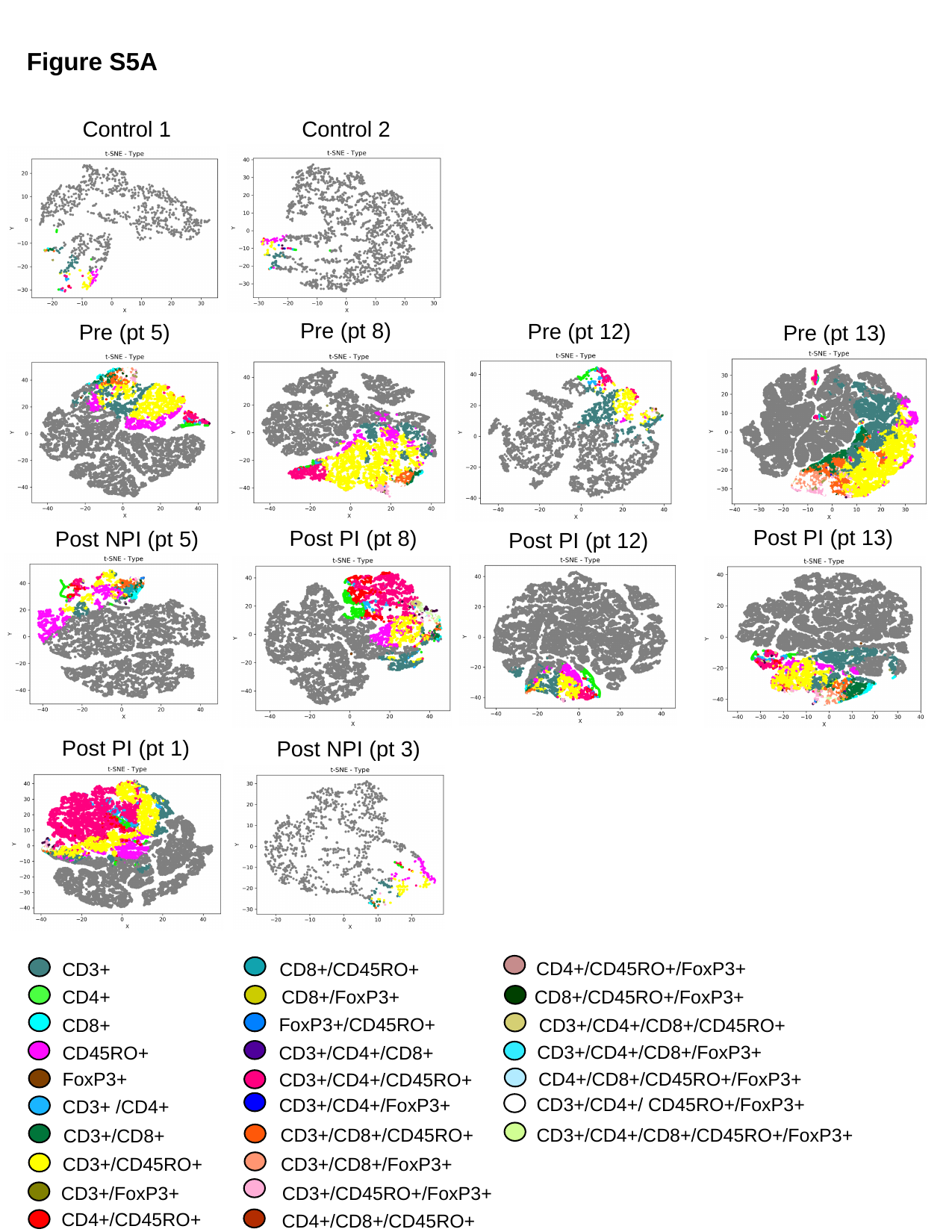

Figure S5A
Control 1
Control 2
Pre (pt 8)
Pre (pt 12)
Pre (pt 5)
Pre (pt 13)
Post PI (pt 13)
Post PI (pt 8)
Post NPI (pt 5)
Post PI (pt 12)
Post PI (pt 1)
Post NPI (pt 3)
CD4+/CD45RO+/FoxP3+
CD8+/CD45RO+
CD3+
CD4+
CD8+/FoxP3+
CD8+/CD45RO+/FoxP3+
FoxP3+/CD45RO+
CD8+
CD3+/CD4+/CD8+/CD45RO+
CD3+/CD4+/CD8+/FoxP3+
CD45RO+
CD3+/CD4+/CD8+
FoxP3+
CD4+/CD8+/CD45RO+/FoxP3+
CD3+/CD4+/CD45RO+
CD3+/CD4+/ CD45RO+/FoxP3+
CD3+/CD4+/FoxP3+
CD3+ /CD4+
CD3+/CD8+/CD45RO+
CD3+/CD4+/CD8+/CD45RO+/FoxP3+
CD3+/CD8+
CD3+/CD8+/FoxP3+
CD3+/CD45RO+
CD3+/CD45RO+/FoxP3+
CD3+/FoxP3+
CD4+/CD45RO+
CD4+/CD8+/CD45RO+

### Slide 6
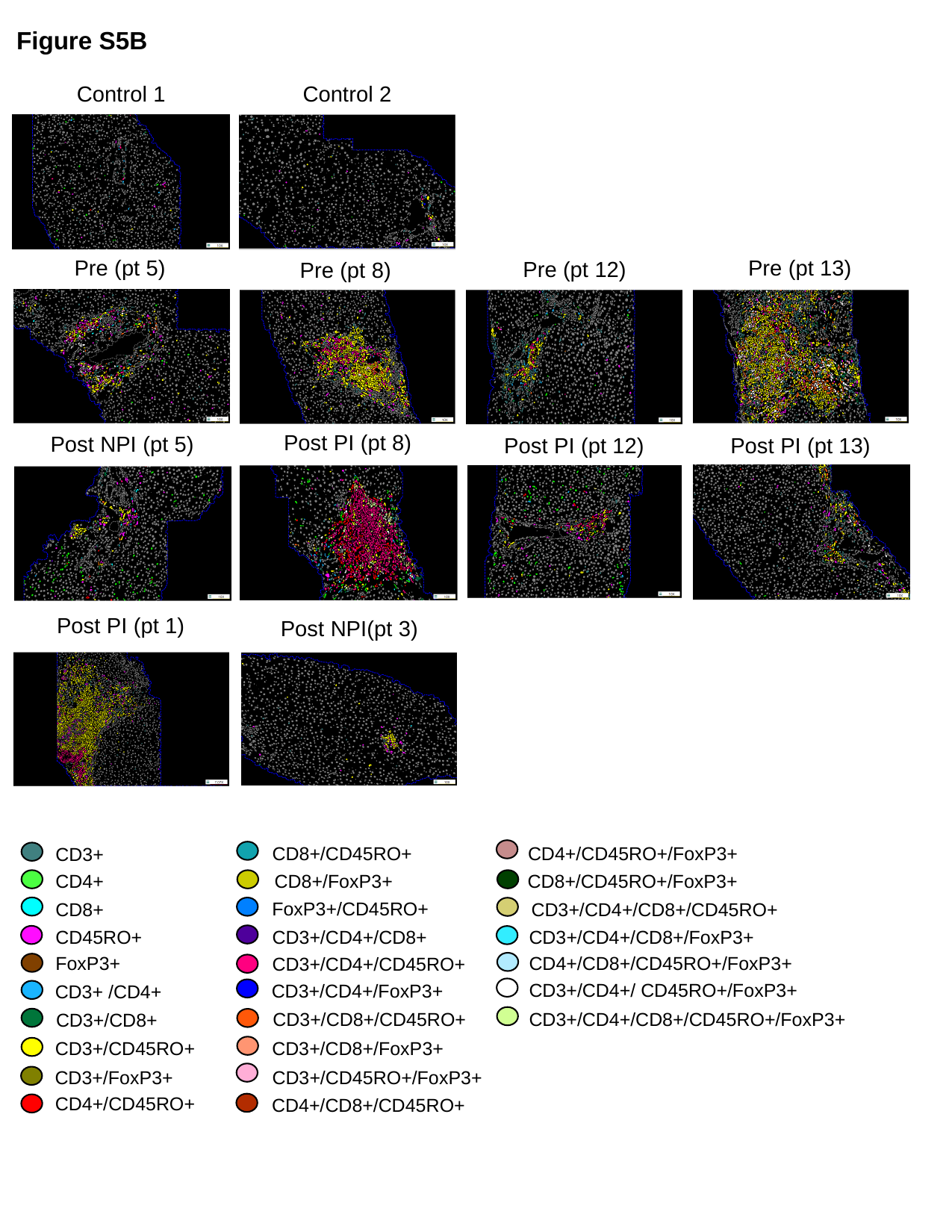

Figure S5B
Control 1
Control 2
Pre (pt 13)
Pre (pt 5)
Pre (pt 12)
Pre (pt 8)
Post PI (pt 8)
Post NPI (pt 5)
Post PI (pt 13)
Post PI (pt 12)
Post PI (pt 1)
Post NPI(pt 3)
CD4+/CD45RO+/FoxP3+
CD8+/CD45RO+
CD3+
CD4+
CD8+/FoxP3+
CD8+/CD45RO+/FoxP3+
FoxP3+/CD45RO+
CD8+
CD3+/CD4+/CD8+/CD45RO+
CD3+/CD4+/CD8+/FoxP3+
CD3+/CD4+/CD8+
CD45RO+
FoxP3+
CD4+/CD8+/CD45RO+/FoxP3+
CD3+/CD4+/CD45RO+
CD3+/CD4+/ CD45RO+/FoxP3+
CD3+/CD4+/FoxP3+
CD3+ /CD4+
CD3+/CD8+/CD45RO+
CD3+/CD4+/CD8+/CD45RO+/FoxP3+
CD3+/CD8+
CD3+/CD8+/FoxP3+
CD3+/CD45RO+
CD3+/CD45RO+/FoxP3+
CD3+/FoxP3+
CD4+/CD45RO+
CD4+/CD8+/CD45RO+

### Slide 7
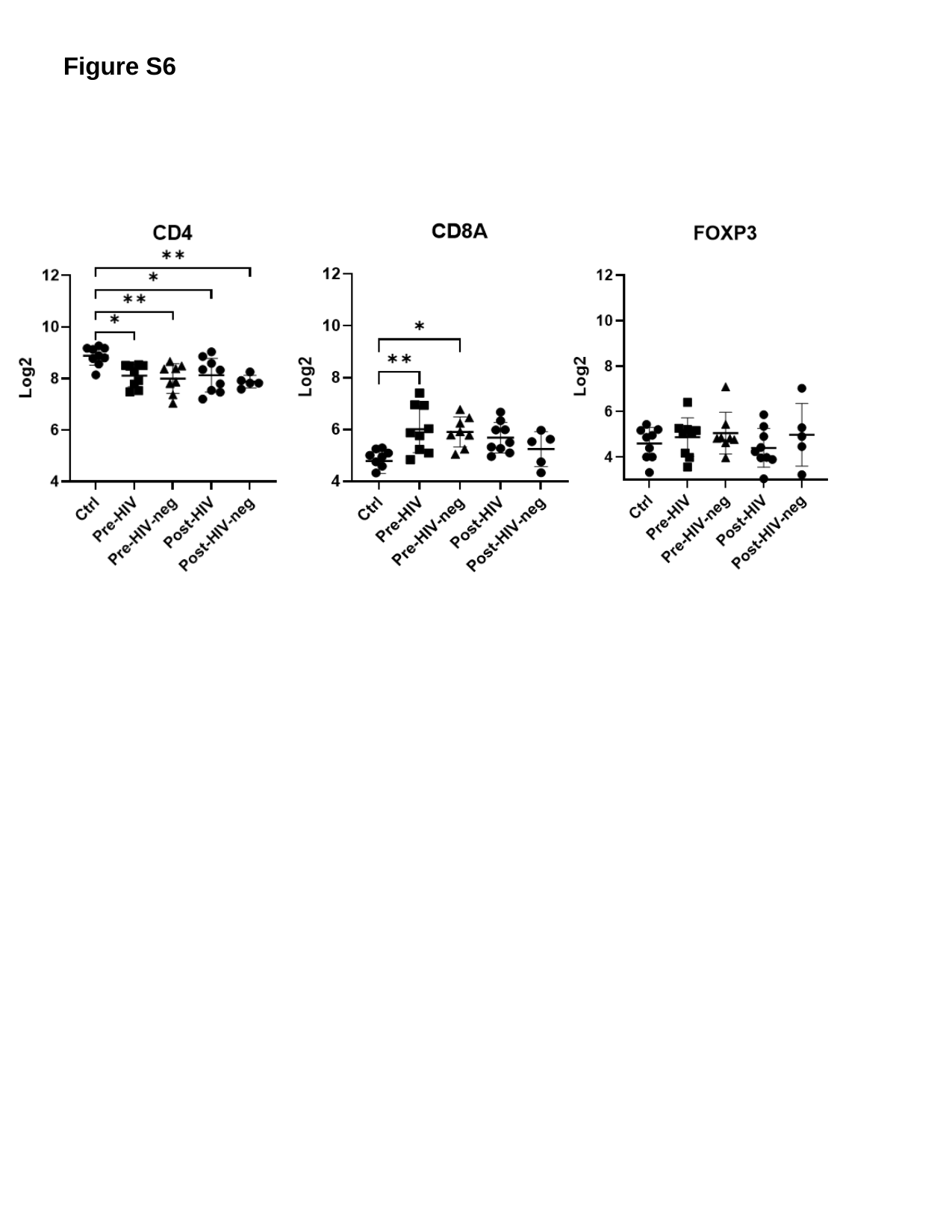

Figure S6

### Slide 8
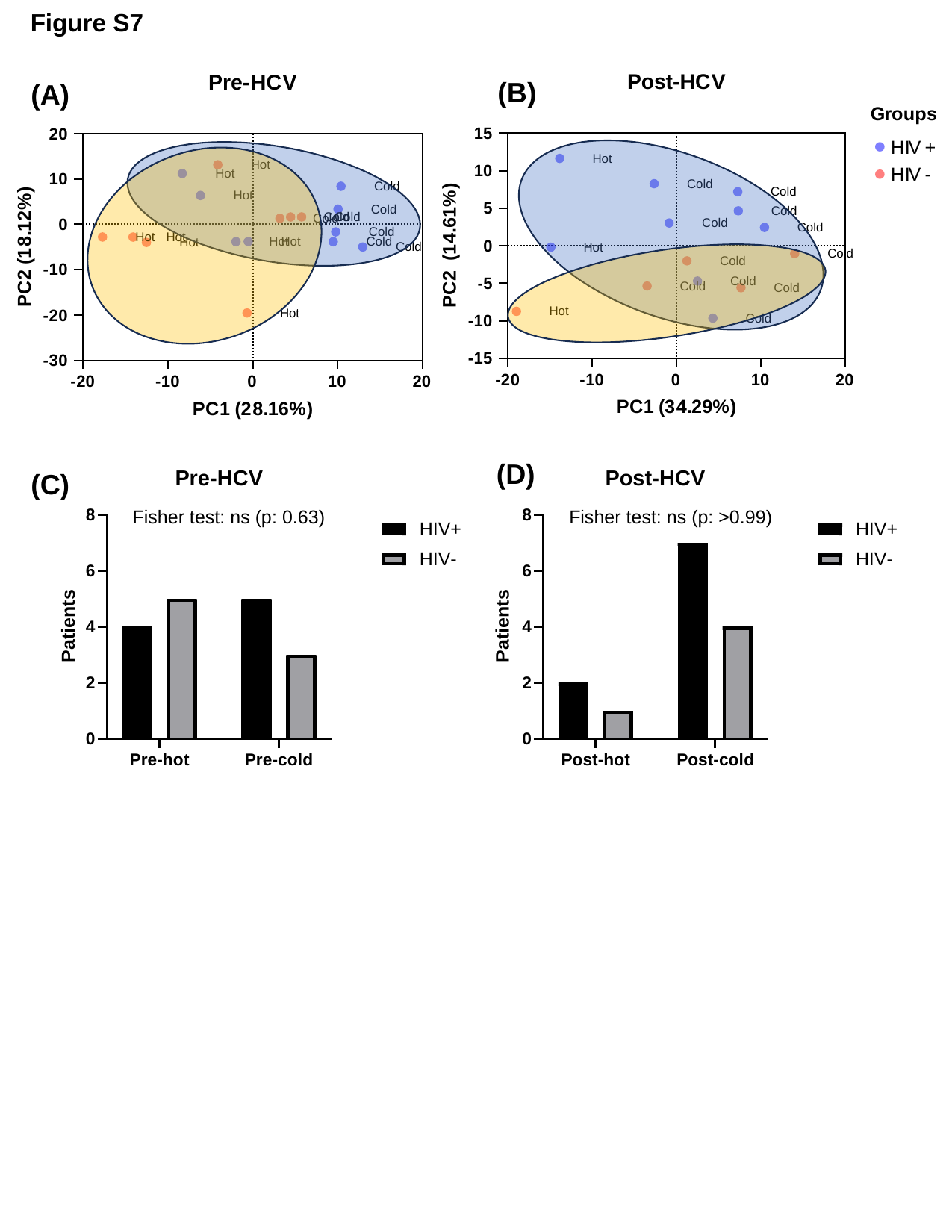

Figure S7
(B)
(A)
(D)
(C)
